# Advanced Care Planning (ACP) in the early phase of COVID-19: A rapid review of the practice and policy lessons learned

**DOI:** 10.1101/2022.09.05.22278731

**Authors:** Sarah Younan, Magnolia Cardona, Ashlyn Sahay, Eileen Willis, Danielle Ni Chroinin

**Author notes:** Corresponding author, Department of Geriatric Medicine, Liverpool Hospital, Elizabeth St, Liverpool, NSW 2170, Australia.

## Abstract

This rapid review of quantitative and qualitative publications of any design indexed in PUBMED between January 2020-April 2021 investigates barriers and enablers of advance care planning (ACP) worldwide in the early stages of the life-threatening COVID-19 pandemic. Seventy-four papers were included: 35 primary research studies (cohorts, reviews, case studies, and cross-sectional designs) and 39 commentaries. Publications from hospitals, outpatient services, aged care and community indicated widespread interest in accelerating ACP documentation to facilitate management decisions and goal-aligned care. Enablers of ACP included targeted public awareness, availability of telehealth, access to online tools and a person-centered approaches. Barriers included uncertainty regarding clinical outcomes, cultural or communication difficulties, legal and ethical considerations, infection control restrictions, lack of time, and limited resources and support systems. The opportunities for rapid implementation of ACP offered by the social distancing restrictions and high demand for health services are valuable in informing future policy and practice.

**What this paper adds:**

- Our study adds to existing evidence by identifying emerging barriers and creative ways of overcoming them in response to a global crisis
- Discussions on death prospects and care of the dying were feasible and a step towards normalisation of advance care planning
- Despite new and overwhelming challenges, policies and practices could be rapidly implemented to satisfy clinicians and families in need of advance care planning

**Applications of study findings:**

- The lessons learnt can be incorporated in future health service planning since the threat of other pandemics is real
- A formal evaluation of effectiveness of some of the emerging strategies would be a valuable addition to the evidence

---

Advance care planning (ACP) has come into sharp focus given the severe impacts of COVID-19 on the old and frail, and pandemic-associated strains on resources (Gupta et al., 2020; Onder et al., 2020; US Centers for Disease Control and Prevention, 2019; Vergano et al., 2020). ACP is “a process that supports adults at any age or stage of health in understanding and sharing their personal values, life goals, and preferences regarding future medical care”, aiming to ensure that medical care received is aligned with values and goals during serious and/or chronic illness (Sudore et al., 2017)

Pre-pandemic, rates of ACP were not high, with population-based estimates of ACP prevalence in Australia of 14% (White et al., 2014) but the US and UK reporting ACP engagement ∼50%, documentation at 33% (Rao et al., 2014; Yadav et al., 2017). Rates of ACP may be lower in population subgroups: black, Asian and minority ethnicity, Lesbian Gay Bisexual Transgender Queer and Intersex, homeless and incarcerated persons and those with limited health literacy (Choi et al., 2020; Ekaireb et al., 2018; Harrison et al., 2016; Hughes & Cartwright, 2015; Kaplan et al., 2020),while disability and female sex may increase ACP uptake (Block, Smith, et al., 2020).

We conducted a rapid review of the literature to explore how the COVID-19 pandemic may have influenced ACP implementation during the early stages of the crisis and investigate barriers and enablers of ACP in the COVID-19 context to inform future policy and practice.

## Method

Given the rapidly evolving nature of the COVID-19 pandemic, we adopted the WHO recommendation of using rapid reviews for the production of actionable evidence (World Health Organization Alliance for Health Policy and Systems Research, 2017).

### Search strategy

We searched PUBMED for the period January 2020-April 2021, using MeSH terms “advance care planning” /“advance directive” plus “COVID-19”/“sars-CoV-2”. We screened reference lists of identified peer-reviewed articles with a focus on ACP during COVID-19.

Inclusion and exclusion criteria. All study designs published were included. Preprints and Non-English language articles were excluded.

### Study selection

SY completed the literature search. MC and SY screened abstracts and reviewed all full texts. DNC, SY and MC reached consensus on all eligibility discrepancies. SY performed data extraction using a pre-defined table (author, publication year, country, study design, publication type, objectives, target population (if applicable), main conclusions). MC cross-checked eligibility for inclusion. Given a rapid review, we did not formally conduct quality appraisal and risk-of-bias assessment.

### Data Synthesis and Reporting

AS and EW reviewed data extraction tables and selected articles. Data were presented using simple descriptive statistics and tables complemented by a narrative synthesis.

## Results

Overall, 343 studies were screened, with 123 full-text papers assessed for eligibility. Afterfull-text screening, 74 were included, comprising commentaries (39) or primary research studies (PRS) (35) (**Figure 1**).

**Figure 1.**
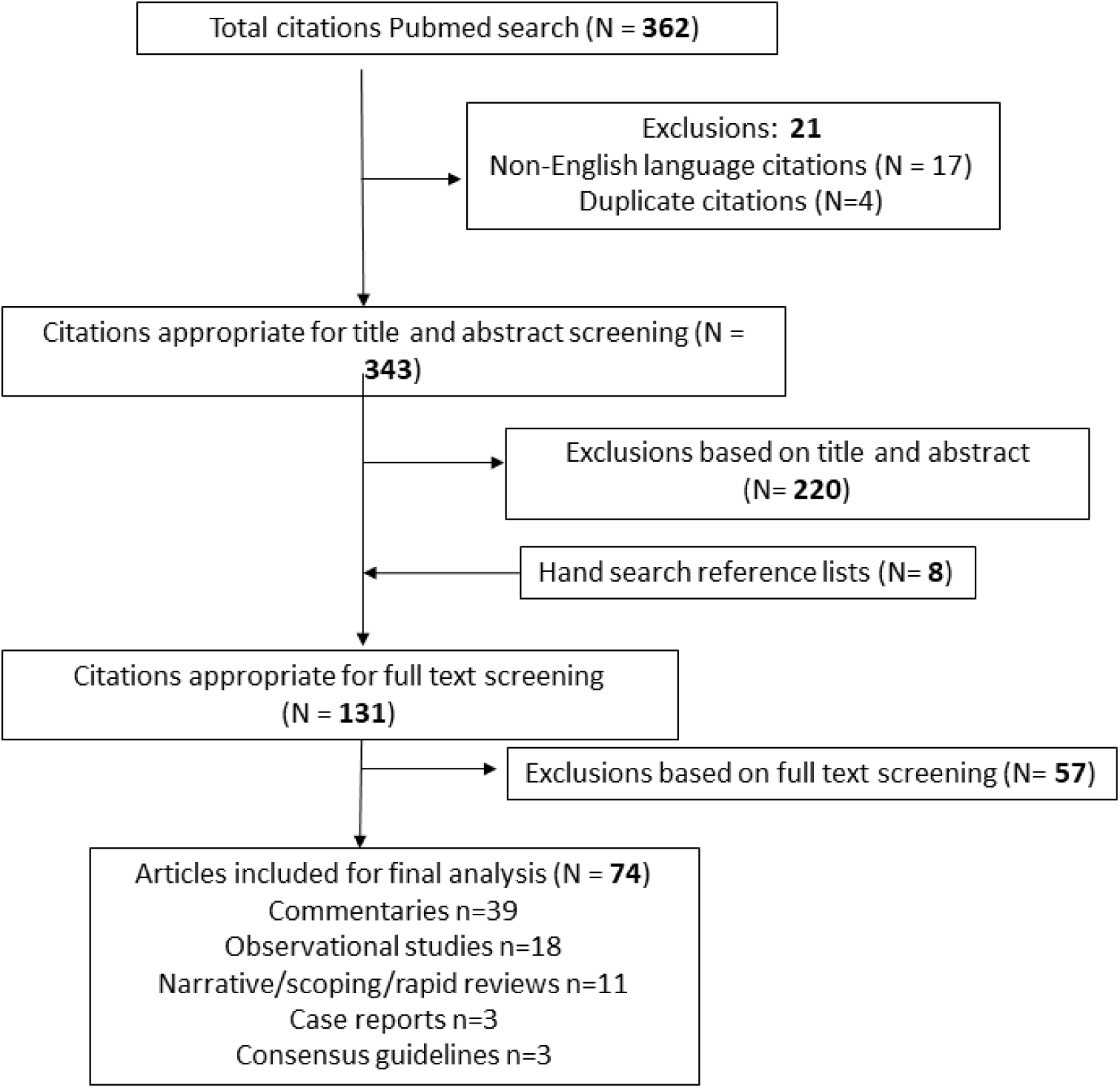
PRISMA diagram of study screening and selection process

Table 1 presents characteristics of included studies; Supplement 1 gives further details. One-third (34%; n=12/35) of the PRS PRS focussed on older adults, the remainder (66%; n=23) targeting mixed age groups.

**Table 1:** Article details for primary studies and commentaries

|  | Primary research<br>(n = 35) | Commentaries<br>(n = 39) |
| --- | --- | --- |
| **Country** |  |  |
| United States of America (USA) | 23 | 26 |
| United Kingdom (UK) | 5 | 5 |
| India | 2 | 1 |
| Taiwan | 2 | 0 |
| Japan | 1 | 0 |
| Netherlands | 1 | 0 |
| South Africa | 1 | 1 |
| Australia | 0 | 2 |
| Canada | 0 | 1 |
| Italy | 0 | 1 |
| New Zealand (NZ) | 0 | 1 |
| Singapore | 0 | 1 |
| **Study Design** | - | 39 |
| Commentaries | 9 |  |
| Cohort study – retrospective | 8 |  |
| Cohort study – prospective | 9 |  |
| Narrative Review | 3 |  |
| Consensus Guidelines | 1 |  |
| Rapid review | 1 |  |
| Scoping Review | 2 |  |
| Case Report | 1 |  |
| Case Series | 1 |  |
| Cross-sectional study |  |  |
| **Study participants/focus** |  |  |
| Patients + Aged Care Clients | 14 |  |
| Policy makers/Guidelines/Ethics | 13 |  |
| General ACP users in the community | 4 |  |
| Clinicians only | 2 |  |
| Mixed: Clinicians + Patients | 2 |  |

There was significant overlap of key discussion points across commentaries and PRS, with themes and sub-themes identified as in Table 2. We explore these themes below.

**Table 2:**
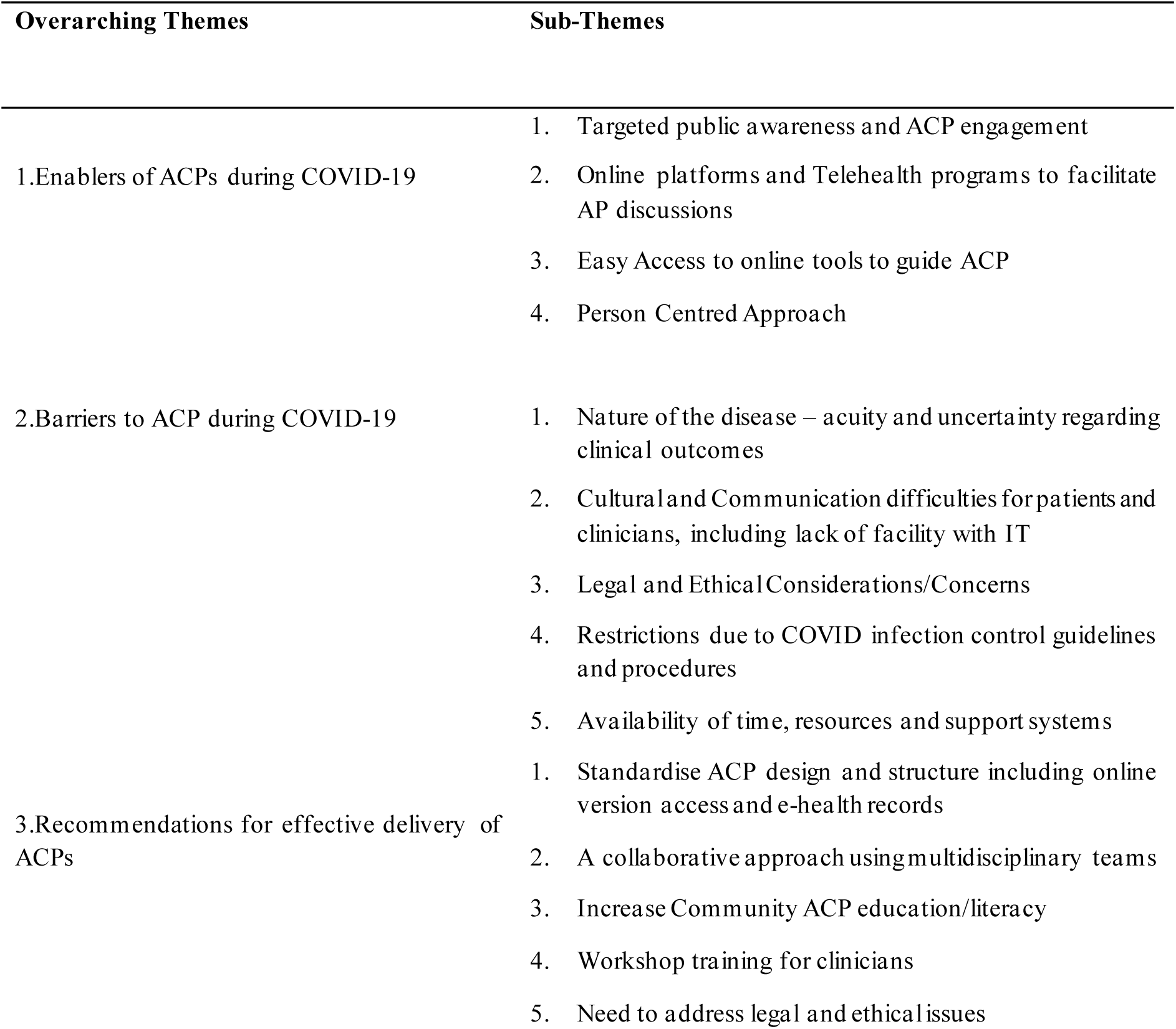
Emerging Themes and Sub-Themes. IT=information technology

| Overarching Themes | Sub-Themes |
| --- | --- |
| 1. Enablers of ACPs during COVID-19 | 1. Targeted public awareness and ACP engagement<br>2. Online platforms and Telehealth programs to facilitate AP discussions<br>3. Easy Access to online tools to guide ACP<br>4. Person Centred Approach |
| 2. Barriers to ACP during COVID-19 | 1. Nature of the disease – acuity and uncertainty regarding clinical outcomes<br>2. Cultural and Communication difficulties for patients and clinicians, including lack of facility with IT<br>3. Legal and Ethical Considerations/Concerns<br>4. Restrictions due to COVID infection control guidelines and procedures<br>5. Availability of time, resources and support systems |
| 3. Recommendations for effective delivery of ACPs | 1. Standardise ACP design and structure including online version access and e-health records<br>2. A collaborative approach using multidisciplinary teams<br>3. Increase Community ACP education/literacy<br>4. Workshop training for clinicians<br>5. Need to address legal and ethical issues |

### Enablers of ACPs during COVID-19

#### Targeted public awareness and ACP engagement

Three studies noted that COVID-19 had increased awareness and importance of completing ACD and legal documentation relating to dying (Morrow-Howell et al., 2020; Portz et al., 2020); one commentator reported a 3.5 fold increase in ACP requests leading to a surge in palliative/supportive care services (Xu et al., 2021). Several commentaries highlighted that the media focus on COVID-19, daily mortality statistics, and stories of dying alone, brought ACP to the forefront of public attention and increased demand for ACPs (McAfee et al., 2020; Sinclair et al., 2020; Xu et al., 2021).

PRS indicated increasing emphasis on ACP for vulnerable groups, such as older adults (Berning et al., 2021; Biswas et al., 2020; Farrell, Ferrante, et al., 2020; Farrell, Francis, et al., 2020; Gilissen et al., 2020; Kotze & Roos, 2020; Kuzuya et al., 2020; Rivet et al., 2020; Valeri et al., 2020; Vipperman et al., 2021; West et al., 2021; Ye et al., 2021), and high-risk groupswith pre-existing life-limiting and/or chronic conditions, such as HIV (Nguyen et al., 2021), cancer or organ failure (Biswas et al., 2020; Rivet et al., 2020; Salins et al., 2020), cardiovascular/cerebrovascular disease (Liao et al., 2020) or severe mental illness (Kotze & Roos, 2020). These recommendations were supported by commentaries (Reja et al., 2020) (Kluger et al., 2020), (Prost et al., 2021).

Commentaries noted that in many aged and acute care settings, ACP was recommended for all admissions (Desai et al., 2020; Gordon et al., 2020). Mooted options included sharing the ACP role with non-medical staff-social workers (Singh et al., 2020), nurses, or volunteers (Desai et al., 2020; Powell & Silveira, 2021; Xu et al., 2021). Some PRS highlighted the value of opportunistic ACP discussions, in outpatient clinics, (Lin et al., 2020; Mills et al., 2021) prior to surgery (Rivet et al., 2020) and/or at admission.

#### Online Platforms and Telehealth programs facilitating ACP

Clinicians, patients and families used on-line platforms and telehealth technology to facilitate ACP. Several PRS and commentaries reported on uptake of outreach telehealth services and/or tool development (Bender et al., 2021; Biswas et al., 2020; DeFilippis et al., 2020; Galbadage et al., 2020; Ho & Neo, 2021; Kuntz et al., 2020; Osterman et al., 2021; Reja et al., 2020).

Telehealth enabled socially/geographically isolated COVID-19 patientsand carers to maintain contact with healthcare professionals (HCPs), engage in ACP conversations, seek psychological support and symptom management (Biswas et al., 2020; Kuntz et al., 2020; Moorman et al., 2020; Osterman et al., 2021). Telehealth also potentially offered benefits for those in financial difficulties (Osterman et al., 2021), facilitated family e-meetings (Kuntz et al., 2020; Piscitello et al., 2021) and enabled carers to engage in virtual farewells with dying patients (Bender et al., 2021; Chan et al., 2021; Dattolo et al., 2021; DeFilippis et al., 2020; Galbadage et al., 2020; Ho & Neo, 2021; Reja et al., 2020). Patients were reportedly relaxed in their home and more willing to engage in ACP discussions (Mills et al., 2021).

Telehealth communication reportedly increased satisfaction rates for clinicians and families (Kuntz et al., 2020). Telehealth aided ACP upskilling and education for clinicians. While clinician ability to initiate ACPs appeared unchanged, one study noted junior doctors’ confidence decreased and anxiety in undertaking ACP rose, possibly a Kruger Dunning effect, where increasing skill acquisition may increase awareness of incompetence (Mills et al., 2021).

#### Easy Access to online tools to guide ACP

Both PRS and commentaries noted accessible online/web-based ACP tools facilitated the development or adaption of COVID-specific end-of-life resources (Auriemma et al., 2020; Gupta et al., 2021; Kluger et al., 2020; Morrow-Howell et al., 2020). Existing tools such as Serious Illness Conversation guide (Paladino et al., 2021) and online workshops (Rabow et al., 2021; Smith et al., 2020) were adapted to context. Online users were relatively young (mean 48 years) and largely female (67%) (Portz et al., 2020).A narrative review (Gupta et al., 2021) also suggested that patient access to online health records facilitates ACP documentation.

#### Person-centred approach

Proponents of ACP argued that effective end-of-life care planning increased compliance with patient wishes and reduced depression and anxiety among bereaved relatives, respects human rights, prevents unnecessary hospital admissions and associated risk of dying alone (Desai et al., 2020; Ho & Neo, 2021; Sinclair et al., 2020).

Several narrative reviews suggested ACP should embrace a person-centred approach to care, involving ‘‘effective’’ or ‘‘adequate, sympathetic’’ communication (Gilissen et al., 2020), and culturally appropriate (West et al., 2021), respecting patients’ autonomy and reflecting their values, choices, and preferences (Farrell, Ferrante, et al., 2020; Gilissen et al., 2020; Rivet et al., 2020; West et al., 2021).

Studies highlighted the importance of identifying a patient’s surrogate decision-maker in emergency situations (Farrell, Ferrante, et al., 2020; Selman et al., 2020). A narrative review cautioned that firstly, critically ill COVID-19 patients may be ill-disposed to make their wishes known, and should not be pressured to make decisions based on conserving resources, and secondly, physicians should not pre-emptively ration or pressurise older adults to reconsider their ACP or resuscitation preferences due resource availability (Farrell, Ferrante, et al., 2020).

### Barriers to ACP during COVID-19

#### Nature of the disease

Both PRS and commentaries identified that the rapid evolution of COVID-19 and uncertain clinical outcomes impeded ACP (Adams, 2020; Back et al., 2020; Ballantyne et al., 2020; Bender et al., 2021; Galbadage et al., 2020; Heyland, 2020; Janssen et al., 2020; Raftery et al., 2020) A lack of prognostic clarity and varied treatment responses delayed ACP (Janssen et al., 2020). Consensus guidelines, by the European Respiratory Society, reported that patients were too ill or anxious to participate in ACP, exacerbated by family absence. Rapid patient deterioration and/or clinical recovery added further complexity to timely discussions about ACPs and/or its currency (Janssen et al., 2020).

#### Cultural and communication difficulties

A narrative review reported that ACP language can becomplex, which may impair communication and lead to inaccurate representation of patient goals (Gupta et al., 2021). Three studies noted that comprehension of ACP forms is influenced by health and literacy levels (Janssen et al., 2020; Lin et al., 2020; Selman et al., 2020), supported by several commentaries (Block, Smith, et al., 2020; Hughes & Vernon, 2021; McAfee et al., 2020; Paladino et al., 2021; Wallace et al., 2020; Xu et al., 2021). A rapid review identified an inverse relationship between ACP and socioeconomic status (SES) (West et al., 2021); with lack of trust in the healthcare system a point echoed in the commentaries (Chan, 2020; West et al., 2021).

Both PRS and commentaries highlighted that ACP uptake is lower in ethnic minorities or lower SES groups (Chan et al., 2021; Gupta et al., 2021; Hughes & Cartwright, 2015; Moore et al., 2020). For example, a narrative review (Gupta et al., 2021) identified that some minority groups may be more likely to appoint a formal ‘Next of Kin’, but less often document ACP, which may increase substitute decision maker burden. Commentary papers highlighted that minority groups lack familiarity with the health system, exacerbated by limited access and communication/language barriers (Block, Jeon, et al., 2020; Hughes & Cartwright, 2015; McAfee et al., 2020; Paladino et al., 2021; Wallace et al., 2020; Xu et al., 2021). Two narrative reviews highlighted potential cultural taboos on discussing death (Gupta et al., 2021; Selman et al., 2020), while a commentary from Sub-Saharan Africa-where ACP is not recognized-noted a focus on efforts to ‘preserve’ life (Essomba et al., 2020). These views may change across generations or time, pointing to the importance of an individualised, patient-centred approach.

Families may also experience communication difficulties. Several commentators reported on family values, perceptions and understandings that differ from those of clinicians (Ballantyne et al., 2020; Chan et al., 2021; Chan,2020; Fausto et al., 2020; Hopkins et al., 2020; Moore et al., 2020).

Clinicians were reminded that familial conflict may compound these differences (Moorman et al., 2020), and that individual preferences can change over time (Janwadkar & Bibler, 2020; Rosenberg et al., 2020). A US retrospective cohort study reported that families were more likely to change life support preferences when they attended in-person bed-side meetings, due to improved understanding of the patient’s condition/prognosis (Piscitello et al., 2021). Commentators also noted that not all population groups have ready access to, or facility with, communication technologies such as telehealth, which may be affected by language or SES (Hughes & Cartwright, 2015; Moore et al., 2020).

Clinicians also experience barriers, for example lack of confidence in communicating bad news (Gupta et al., 2021) and difficulties in making speech heard when wearing personal protective equipment (PPE) (Selman et al., 2020). These barriers ranged from difficulties initiating ACP discussions with patients/families (Gupta et al., 2021; Selman et al., 2020) to concerns surrounding topic sensitivity and family grief (Janssen et al., 2020; Lin et al., 2020; Selman et al., 2020). For example, clinicians may delay end-of-life conversations, reluctant to ‘take away’ hope when prognosis is uncertain. A European Taskforce suggested a constant review process will maintain alignment of ACP to the patient’s evolving condition (Janssen et al., 2020). Three PRS and several commentaries recommended the need for clinician ACP training and access to palliative care education (Gupta et al., 2021; Janssen et al., 2020; Selman et al., 2020). Mitigating strategies proposed included ACP integration at all training levels (McAfee et al., 2020), specific frameworks such as VitalTalk (Back et al., 2020) or other conversation guides to ensure equitable access and goal-aligned management (Desai et al., 2020; Essomba et al., 2020; Prost et al., 2021), and ACP upskilling for allied health providers (Raftery et al., 2020).

#### Legal and Ethical Considerations

ACP laws and guidelines differ across countries and jurisdictions. Difficulties range from accessing lawyers (Chan, 2020), to differing legal terminology (Block, Smith, et al., 2020), to identifying who is entitled to initiate conversations. In some jurisdictions, only medical staff may initiate ACP conversations, with limited/no input from other multidisciplinary team members (Raftery et al., 2020; Singh et al., 2021). A large cohort study conducted in the US, found engaging social workers in ACP resulted in a 13% increase in patients nominating a medical POA (Singh et al., 2021).. A Taiwanese study noted local legal obligations to have two witnesses and associated costs might impede ACP (Lin et al., 2021). A narrative review during the pandemic (Gilissen et al., 2020) identified seven documents that described core ACP issues to be discussed with nursing home residents: POA (Enduring Guardian equivalent), cardiopulmonary resuscitation (CPR), admission to hospital, intubation, non-invasive ventilation, fluids, antibiotics, etc. Supplement 1 includes related resources, including a position statement.

A Canadian commentary argued that ACPs are less useful for COVID-19, lacking situational value, and difficulty fully informing individuals regarding treatment options, benefits and risks (Heyland, 2020). They suggest patients should rather be asked about values, noting that ethical and legal issues are of serious concern during a pandemic. Another commentary reported that some standardized protocols, e.g. in the UK, suggested that residential aged-care residents should not be admitted to hospitals (Iacobucci, 2020). Such “one-size fits-all” approaches contradict the uniqueness of individual presentations (Iacobucci, 2020).

A US consensus guideline highlighted encouraged decision makers to focus on short-term outcomes and avoid age alone as a determinant of care. Other recommendations included avoiding ancillary criteria such as predicted long-term life expectancy (disadvantages elderly), formation of triage committees and development of transparent resource-allocation strategies to facilitate appropriate ACP (Farrell, Ferrante, et al., 2020).

#### Restrictions due to COVID infection control procedures

Almost all studies identified that COVID-related restrictions impacted peoples’ ability to connect with clinicians and access ACP-related support. Infection control protocols governing the spread of COVID (social distancing, mask-wearing) impacted on communication between patients, families and HCPs. This, in turn, decreased the uptake of ACPs as outlined in PRS (Coleman et al., 2020; Janssen et al., 2020; Lin et al., 2021) and commentaries (Gordon et al., 2020; Paladino et al., 2021). For example, a UK based cohort study identified a decline (from 75.4% to 50.6%) in Do-Not-Attempt-Cardiopulmonary-Resuscitation due to restricted family visiting (Coleman et al., 2020). The use of PPE and restricted visiting were hurdles to building trust required for effective ACP discussions (Mills et al., 2021); (Janssen et al., 2020). These findings were ratified overwhelmingly by the commentaries (Adams, 2020; Ballantyne et al., 2020; Bender et al., 2021; Block, Jeon, et al., 2020; Chan et al., 2021; Ho & Neo, 2021; Hopkins et al., 2020; Janwadkar & Bibler, 2020; Moorman et al., 2020; Wallace et al., 2020; Xu et al., 2021).

Lack of interpersonal access increased family and patient distrust/suspicion towards HCPs (Moore et al., 2020), and/or the healthcare system (Gordon et al., 2020), [85]. Isolation, travel restrictions, and visitor limitations made shared decision-making extremely difficult. This was seen from the potentially-dying patient deprived of in-person visits (Adams, 2020; Ballantyne et al., 2020; Bender et al., 2021; Galbadage et al., 2020; Gordon et al., 2020; Moorman et al., 2020; Paladino et al., 2021; Wallace et al., 2020), to feelings of being abandoned by medical staff (Janwadkar & Bibler, 2020). Stigma surrounding COVID, reinforced by the infection control protocols, exacerbated these difficulties and created barriers to ACP (Essomba et al., 2020).

#### Lack of time, resources and support systems

ACP was identified as enablingfairer and more transparent resource allocation during periods of constraint, potentially reducing unnecessary life-sustaining treatments and streamlining the use of intensive care resources (Farrell, Francis, et al., 2020; Gupta et al., 2020; Liao et al., 2020; Salins et al., 2020). This was seen to alleviate the need for healthcare rationing, by avoiding aggressive treatment for those who do not want it, engaging patients in shared-decision making and avoiding blanket decisions based on age or comorbidity. Four commentaries noted that ACP also reduced clinician exposure to avoidable infection and risk (e.g. with CPR), and assisted in planning for surges in healthcare demand (Ho & Neo, 2021; Rosenberg et al., 2020; Sinclair et al., 2020; Xu et al., 2021).

Nonetheless, ACP was recognised to be time intensive. Several PRS highlighted that clinicians are time poor, with little time for lengthy discussions or to develop clinician-patient relationships and trust (Gilissen et al., 2020; Gupta et al., 2021; Liao et al., 2020; Salins et al., 2020; Selman et al., 2020). Numerous commentaries supported this finding (Ballantyne et al., 2020; Bender et al., 2021; Janwadkar & Bibler, 2020; Moore et al., 2020; Paladino et al., 2021; Raftery et al., 2020; Wallace et al., 2020). Commentators noted a lack of appropriate policies highlighting ACP’s importance, a failure to prioritize it (Adams, 2020; Back et al., 2020; Bender et al., 2021; Chan et al., 2021; Ho & Neo, 2021), and either poor or overly-cumbersome documentation, as well as failure to remunerate appropriately e.g. with dedicated billing codes(Bender et al., 2021).

Requests for ACP have reportedly increased since the pandemic across varied settings: aged-care, terminal neurological patients, and prisons (Bolt et al., 2021; Desai et al., 2020; Gordon et al., 2020; Kluger et al., 2020; Lin et al., 2020; Morrow-Howell et al., 2020; Prost et al., 2021; West et al., 2021), with goals of care discussions ranging from 36% (Piscitello et al., 2021) to a 3.5-fold increase (Xu et al., 2021).

### Recommendations for effective ACP delivery

#### ACP design and structure

Various commentators suggested a need to develop or adapt tools specifically to the COVID context (Kluger et al., 2020; Morrow-Howell et al., 2020), and to deliver small chunks of information given the emotional impact of the diagnosis (Back et al., 2020; Block, Smith, et al., 2020; Hopkins et al., 2020; Singh et al., 2020). Three editorials suggested current ACP tools are targeted towards non-communicable chronic illnesses rather than the COVID experience (Ballantyne et al., 2020; Moore et al., 2020; Moorman et al., 2020). Others provided specific ACP tools, approaches or frameworks (Gaur et al., 2020; Paladino et al., 2021; Roberts et al., 2020). Similar suggestions were made in PRS,, but specific recommendations for design and structure of ACPs were limited.

#### A collaborative approach

Multidisciplinary collaboration was encouraged, guided by specialists trained in ACP or palliative care (Berning et al., 2021; Bolt et al., 2021; Powell & Silveira, 2021), although noting that specialist teams are not universally available (Prost et al., 2021). Other suggestions included that non-medical team-members should assist with ACP, e.g. nurses and social workers (Desai et al., 2020; Raftery et al., 2020), or spiritual carers (Xu et al., 2021). Frameworks and tools were seen to address such concerns and to improve clinician confidence (Desai et al., 2020; Prost et al., 2021; Singh et al., 2020). A commentary suggested that interpersonal barriers of social distancing could be overcome by increasing pre-illness ACP discussions (Ballantyne et al., 2020).

#### Increase Community ACP education/literacy

The importance of promoting and increasing ACP education and literacy for the wider community was frequently recognised (Janssen et al., 2020; McAfee et al., 2020; McMahan et al., 2021; Selman et al., 2020; Singh et al., 2020), including relationship-building with community, ethnic and religious groups (Hughes & Vernon, 2021; Xu et al., 2021).

One commentary suggested that hospital-based policies, jurisdictional legislation and community campaigns needed to accompany the broadening ACP education for HCPs (Raftery et al., 2020). Death education/literacy may be best embedded across all health curriculum levels (McAfee et al., 2020). Other options proposed included templates and tools to facilitate ACPs, videos and telehealth to facilitate remote ACP, and creation of an integrated web-based system linked to electronic medical records (eMR) to facilitate ACP in the community (Galbadage et al., 2020; McMahan et al., 2021; Portz et al., 2020; Selman et al., 2020; Smith et al., 2020).

Several other studies indicated a need to promote ACP literacy, but specific strategies were lacking (Auriemma et al., 2020; Funk et al., 2020; Kuzuya et al., 2020; Lin et al., 2021).

#### Training for clinicians

Upskilling clinicians to improve ACP confidence was a recurring theme, highlighted by at least two narrative reviews (Gupta et al., 2021; Selman L et al., 2020) and a consensus guideline (Janssen et al., 2020). Proposed teaching strategies were varied, e.g. role-play, interactive multimedia, virtual training, online communication, and telephone debriefing (McMahan et al., 2021; Mills et al., 2021). One review suggested that training HCPs in assisted living communities could help communication across services (Vipperman et al., 2021).

#### Need to address legal and ethical issues

Legal and ethical issues featured predominantly in commentaries. Suggestions included temporarily pausing stricter legal requirements for ACP during COVID (Block, Smith, et al., 2020). Several studies addressed resource utilization and most studies argued that ACP should be prioritized to reduce unwanted intensive/life-sustaining treatment in a stretched healthcare system (Curtis et al., 2020). Clinician concerns regarding the ethics of CPR in COVID-19 patients were also noted (DeFilippis et al., 2020). Nevertheless, one institution highlighted that resource availability and patient/family goals of care need to be balanced when making resuscitation decisions (Fausto et al., 2020).

## Discussion

This rapid review of ACP implementation in the initial phase of the pandemic (until April 2021) suggests a high level of worldwide interest since COVID emerged. ACP may benefit overwhelmed healthcare systems, and decision-making for patients, caregivers and clinicians, and improve bereavement experiences. A sense of urgency and eagerness to make recommendations was apparent, often without supporting high-quality evidence (39 commentaries, 3 consensus guidelines and 3 case series/reports). Most publications were from the USA and UK ─ severely affected countries but also resourced to produce and promote research. Settings spanning acute hospitals, community, aged care and outpatient services showcased the breadth of the locations affected. We were not surprised by the focus on barriers and enablers of ACP during the pandemic. The variety of proposed solutions was reassuring in terms of timeliness and feasibility, with standardization, collaboration, education and ethical-legal considerations underpinning implementation into practice.

Two main lessons surfaced from this rapid review. First, the prevalence and clarity of goals of care discussions were often touched on as a priority issue for patient management, prevention of unwanted (non-goal-aligned) care, or transfer decisions, across the range of study types ((Bolt et al., 2021; Burke et al., 2020; Gupta et al., 2021; Kuntz et al., 2020; Liao et al., 2020; Piscitello et al., 2021; Salins et al., 2020; Vipperman et al., 2021)(Nguyen et al., 2021). Next, frequent references to online tools or resources (Auriemma et al., 2020; Gaur et al., 2020; Paladino et al., 2021; Portz et al., 2020; Selman L et al., 2020) and virtual learning for clinicians and families including online family meetings (Gupta et al., 2021; Janssen et al., 2020; Kuntz et al., 2020; Mills et al., 2021; Piscitello et al., 2021; Rabow et al., 2021; Smith et al., 2020), reflect rapid embracing of innovative approaches to facilitate ACP completion and circumvent face-to-face interactions. Both are reassuring findings which indicate that patient/family wishes on end-of-life care were not neglected in the chaos of the unanticipated demand for health services, although frequency and equity of access were not always optimal (Gupta et al., 2021; Piscitello et al., 2021), and absence of communication was perceived to lead to complex bereavement (Carr et al., 2020; Selman et al., 2020).

A poignant aspect of COVID-19 was the vision of people dying in alone in hospital, unable to be with family and suffering respiratory discomfort. For families, this led to a complicated grief experience characterized by survivor guilt, anger and distress. An obvious mitigating strategy was to increase ACP so all stakeholders were prepared, patients received the care they desired and symptoms were managed in line with individualised goals (Carr et al., 2020; Cavalier et al., 2020; Desai et al., 2020; Hughes & Vernon, 2021; Janwadkar & Bibler, 2020).

### Implications for practice

Our pandemic-related experience has pushed us to re-evaluate existing healthcare policies and practices, overcome new and longstanding barriers, and embrace new solutions, in ACP as in other areas of practice. This review suggests the pandemic has provided some impetus to drive adaptable ACP conversations at individual, local, and international levels, affording an opportunity for longer term improvements in ACP practice and care that is respectful of, and responsive to, the values and needs of the individual patients we meet every day.

### Limitations of the review

Given the nature of a rapid review, we only used limited number of terms, confined to searching English language publications in the early stages of the pandemic and from a single (but the largest) database anticipated to contain the majority of relevant articles. Half of the publications reflected perception, early reactions and proposed solutions and were not interventions or evaluations of policies or practices. Our intention was to illustrate all perspectives given the uniqueness of this serious pandemic experience. A future review of subsequent publications in the late stages of the review may report similar or different findings and it would be interesting to compare our results with emerging lessons after longer exposure to the global threat.

## Conclusion

Despite high demand for healthcare services, the pandemic provided opportunities for rapid implementation of ACP. Both barriers and enablers of ACP influence its uptake Evidence suggests clinicians, patients and families support the cultural shift that fosters positive ACP uptake; this may contribute to reduced overtreatment near end of life. Ongoing evaluation of policy changes, effectiveness and acceptability are warranted.

## Data Availability

All data produced in the present work are contained in the manuscript

## Data Availability

All data produced in the present work are contained in the manuscript

## Acknowledgments

The authors thank our institutional librarians for their assistance finding full texts.

## Conflict of Interest

The authors declare no financial or other actual or perceived conflicts of interest in this work.

## Funding

This study received no funding from any government, commercial or no-for-profit sources.

## Ethical approval

Not relevant to this methodology.

---

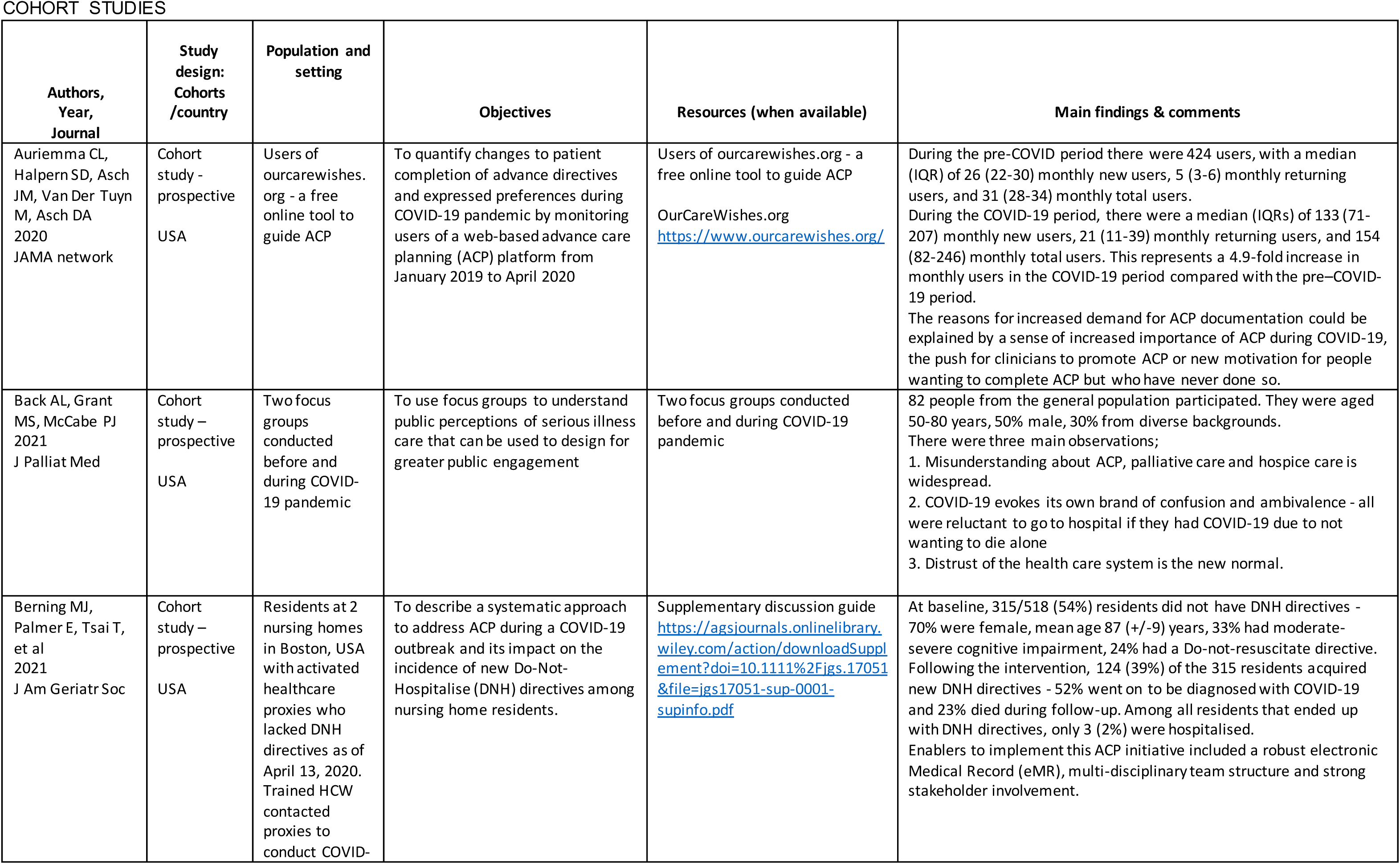
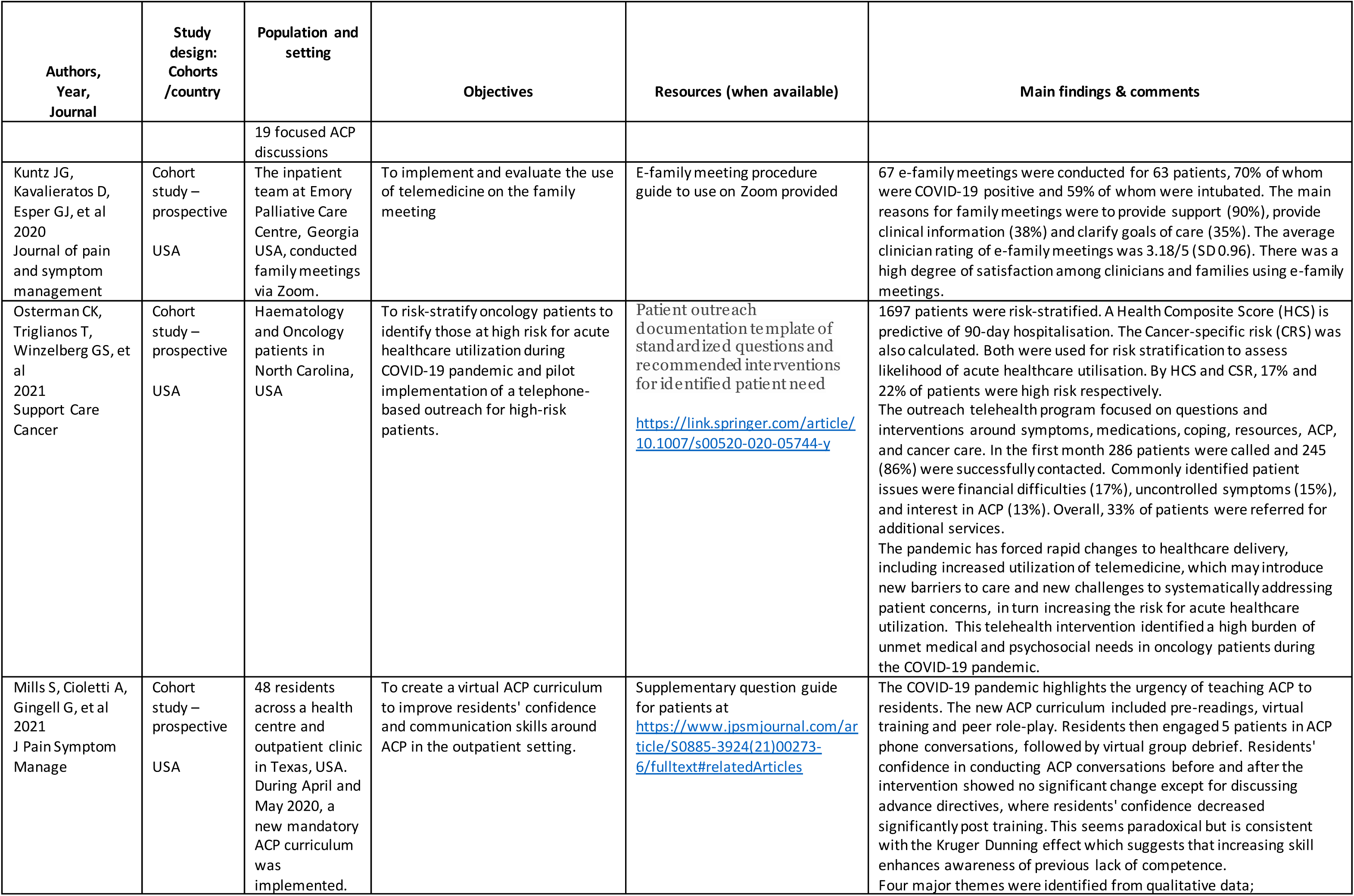
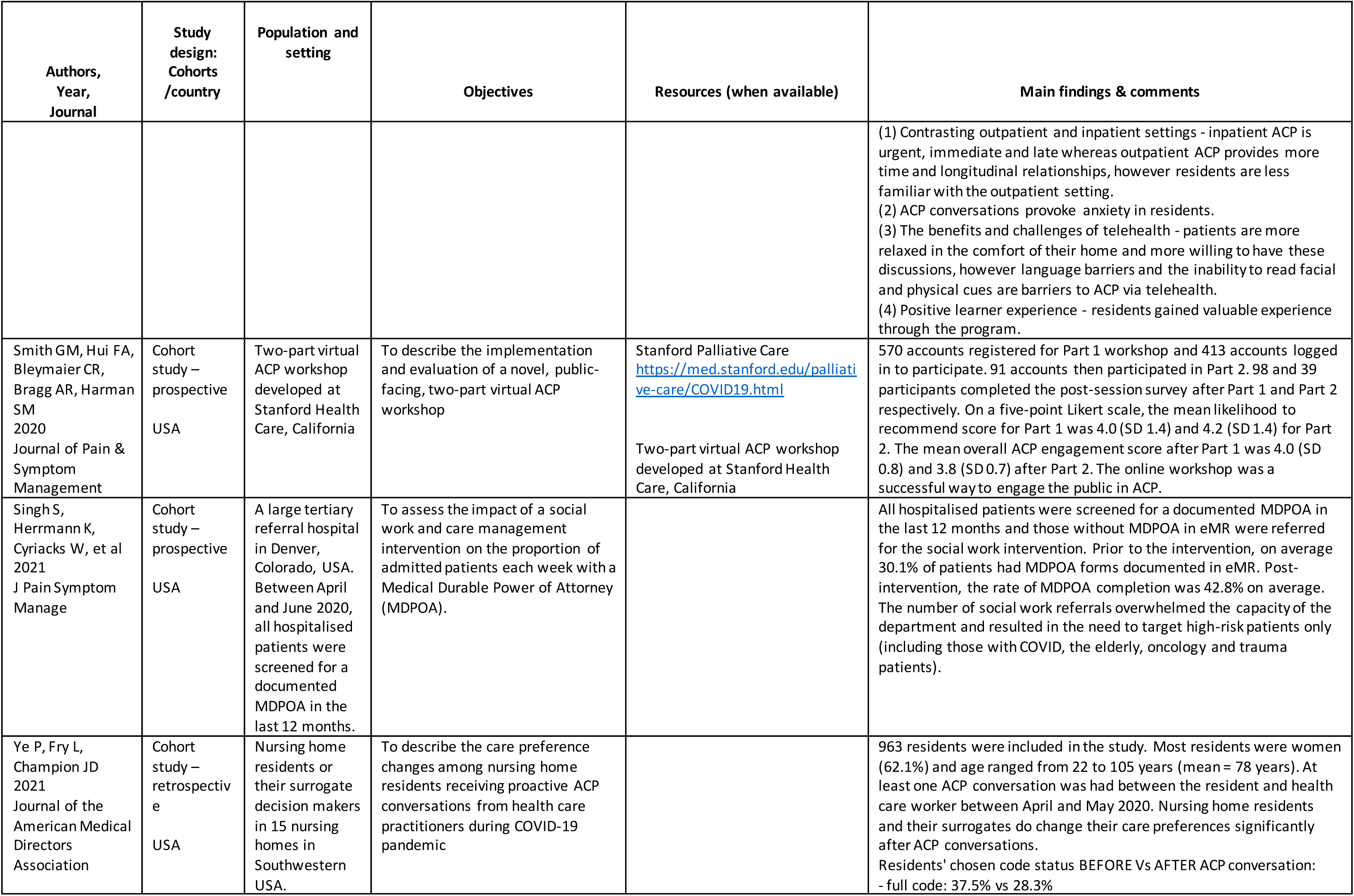
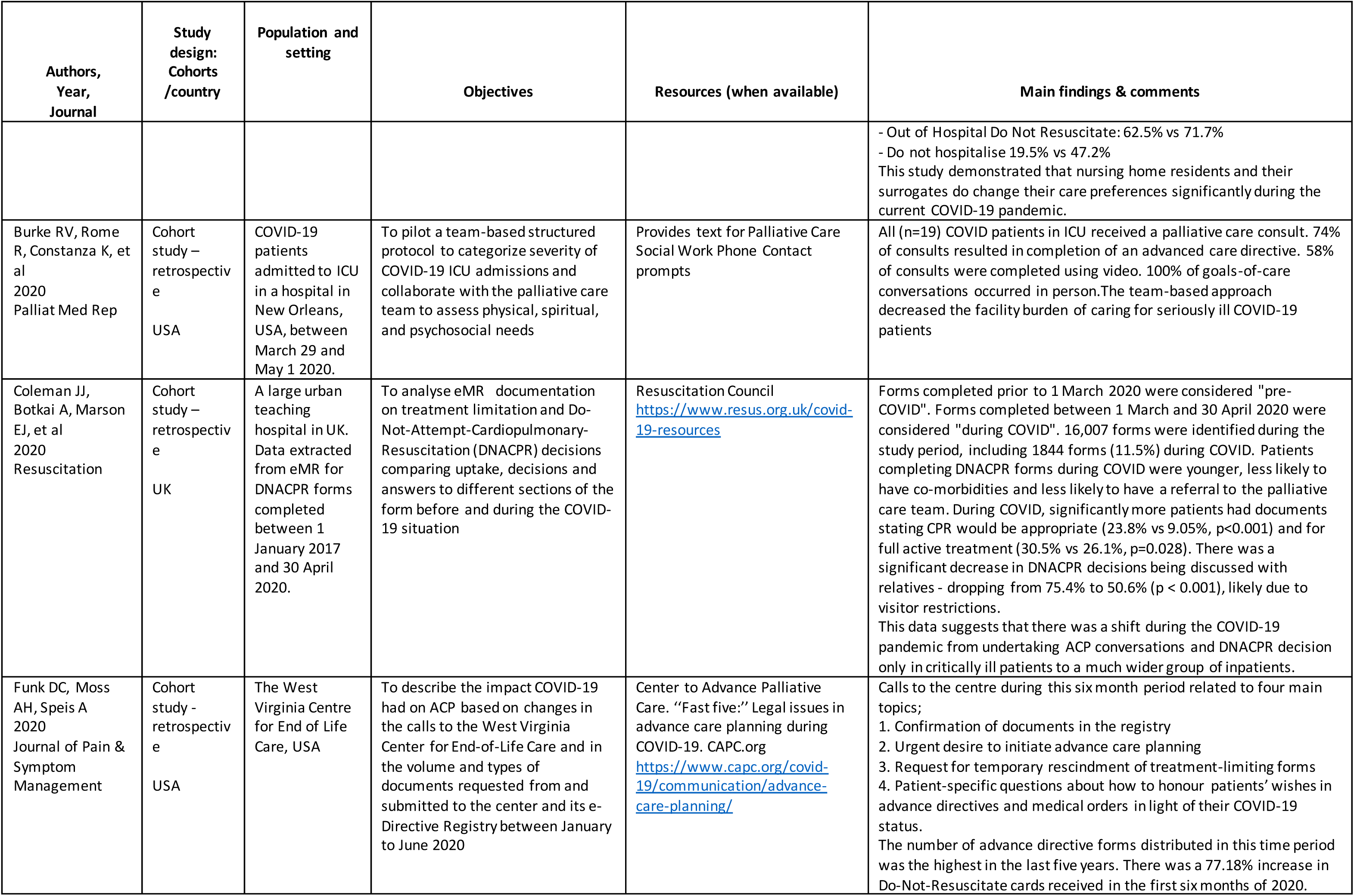
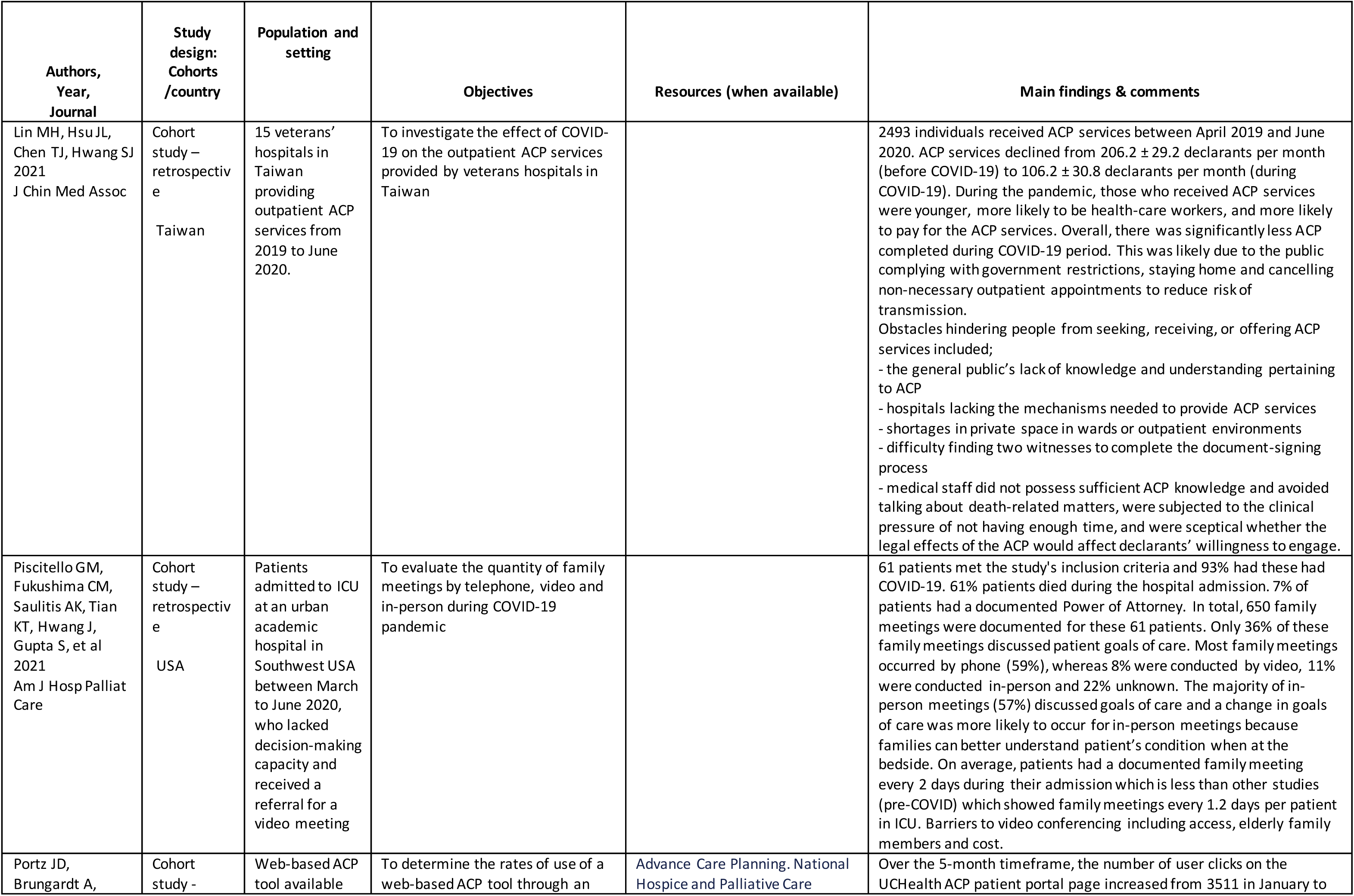
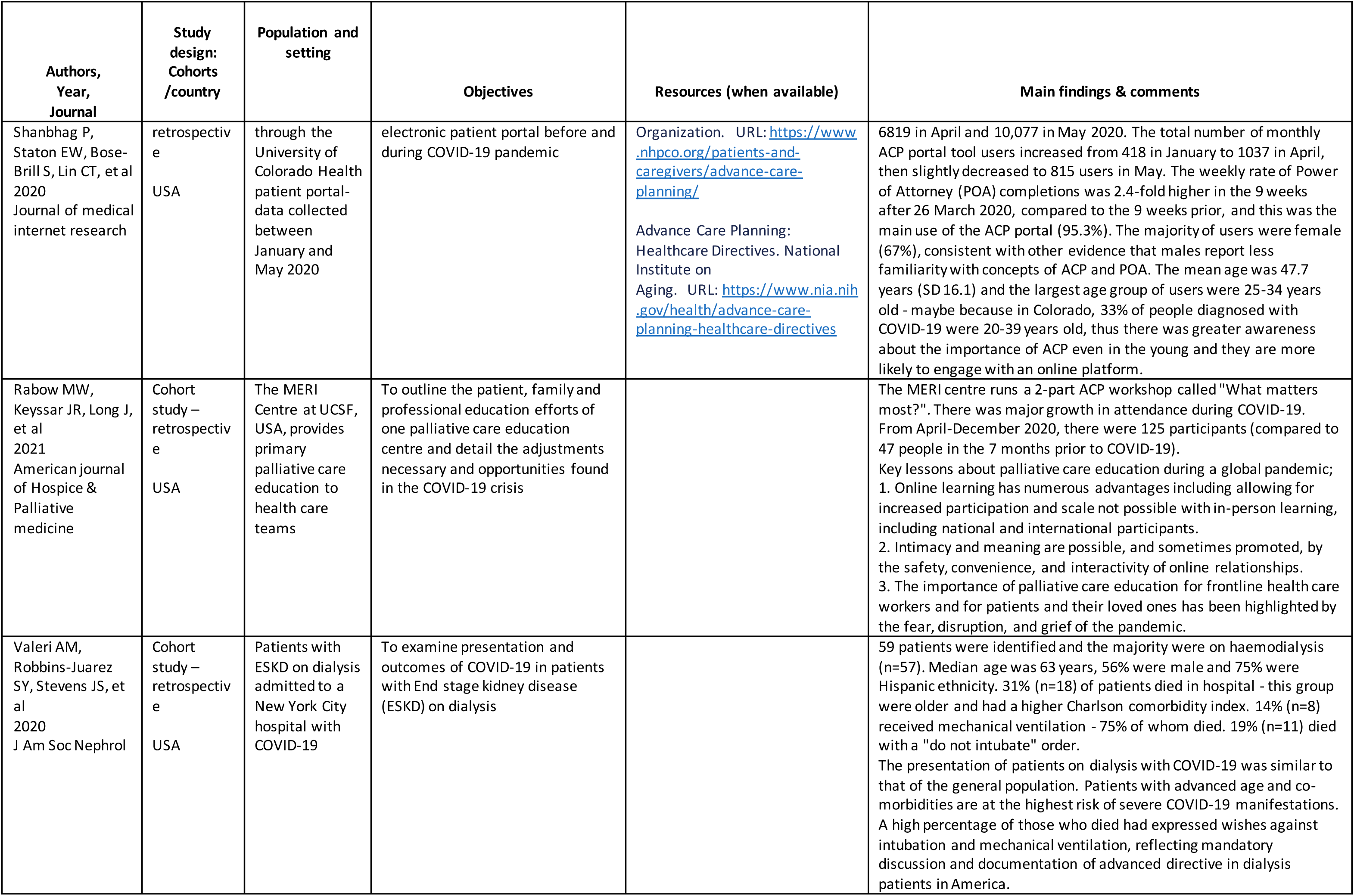
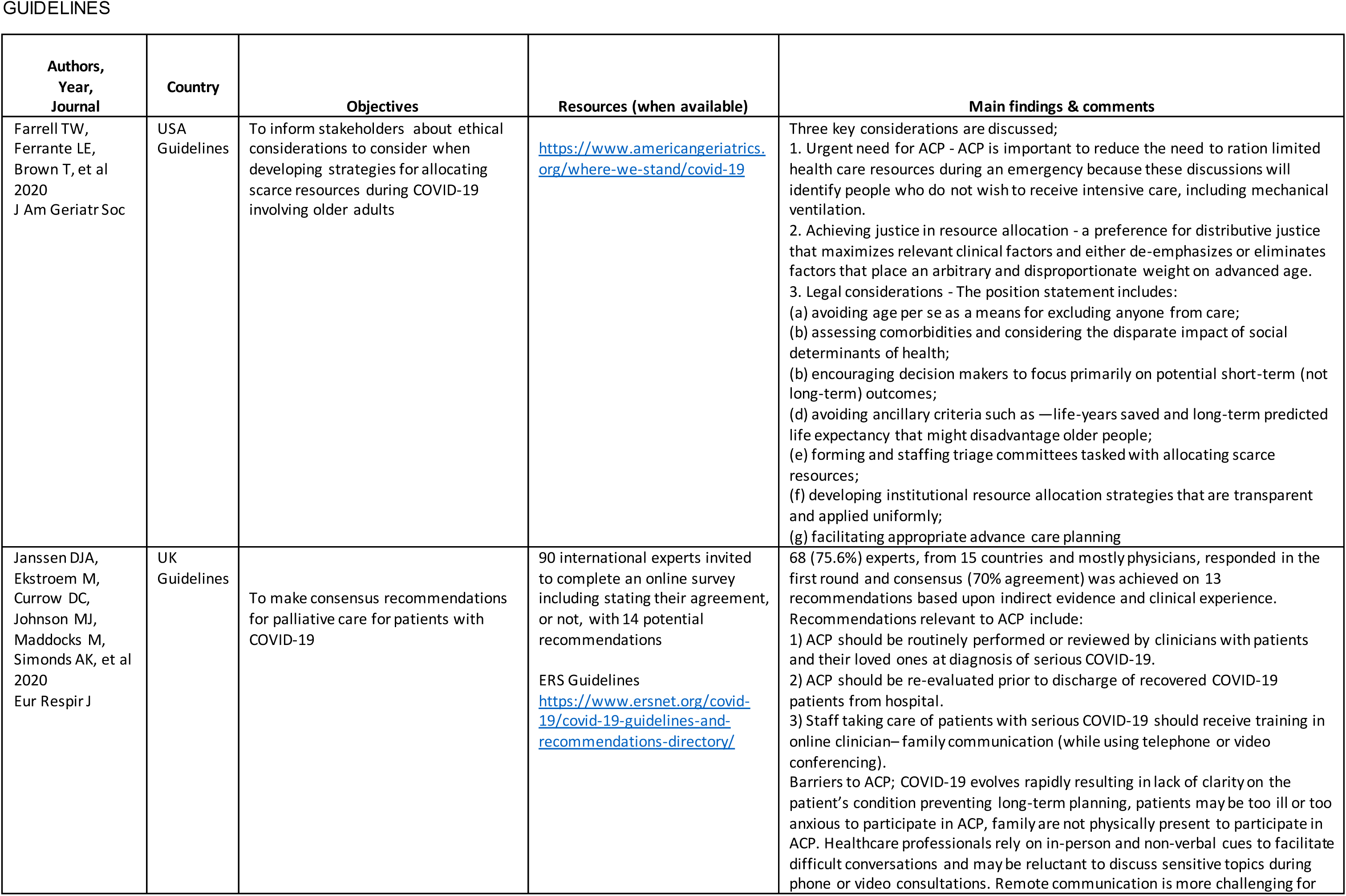
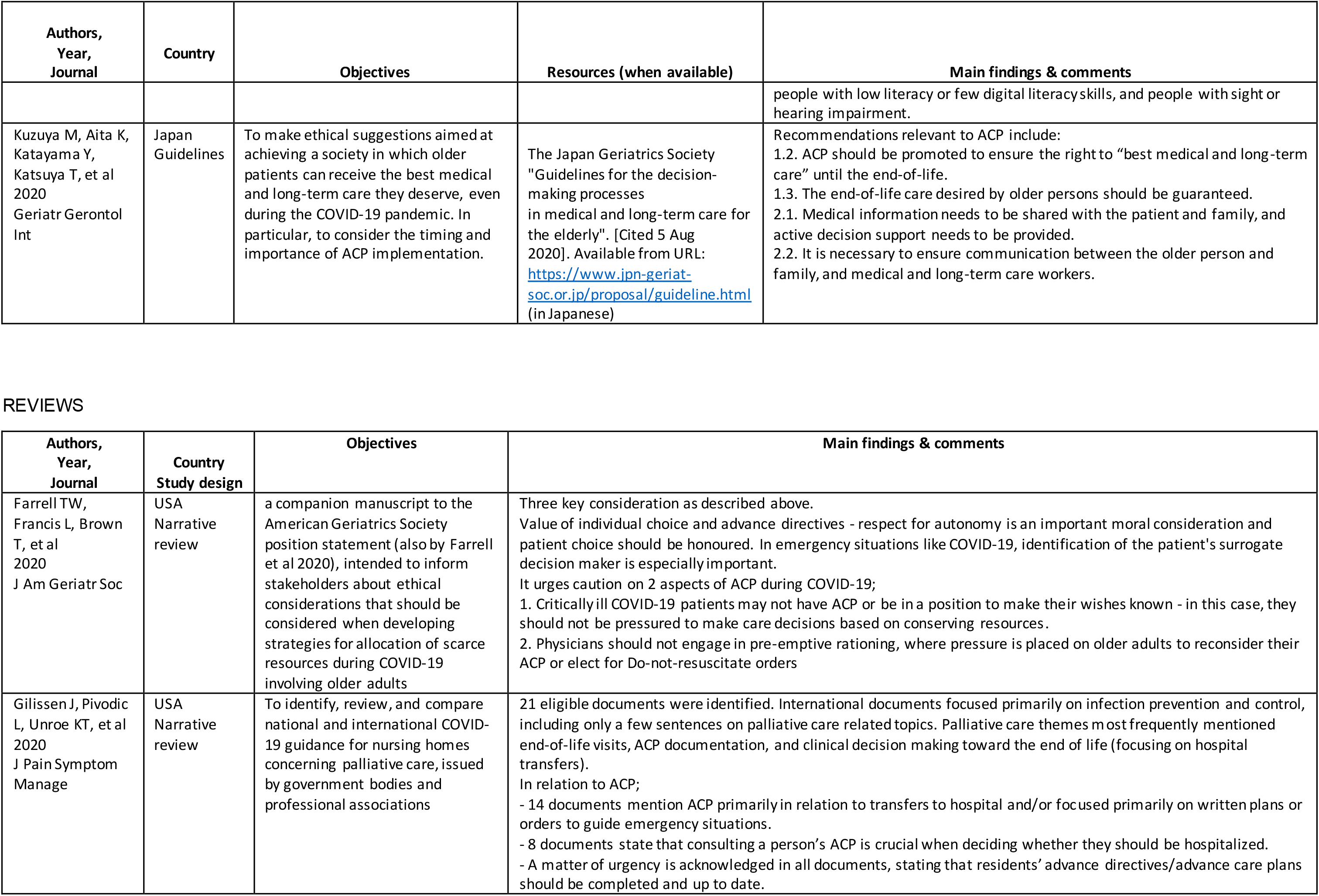
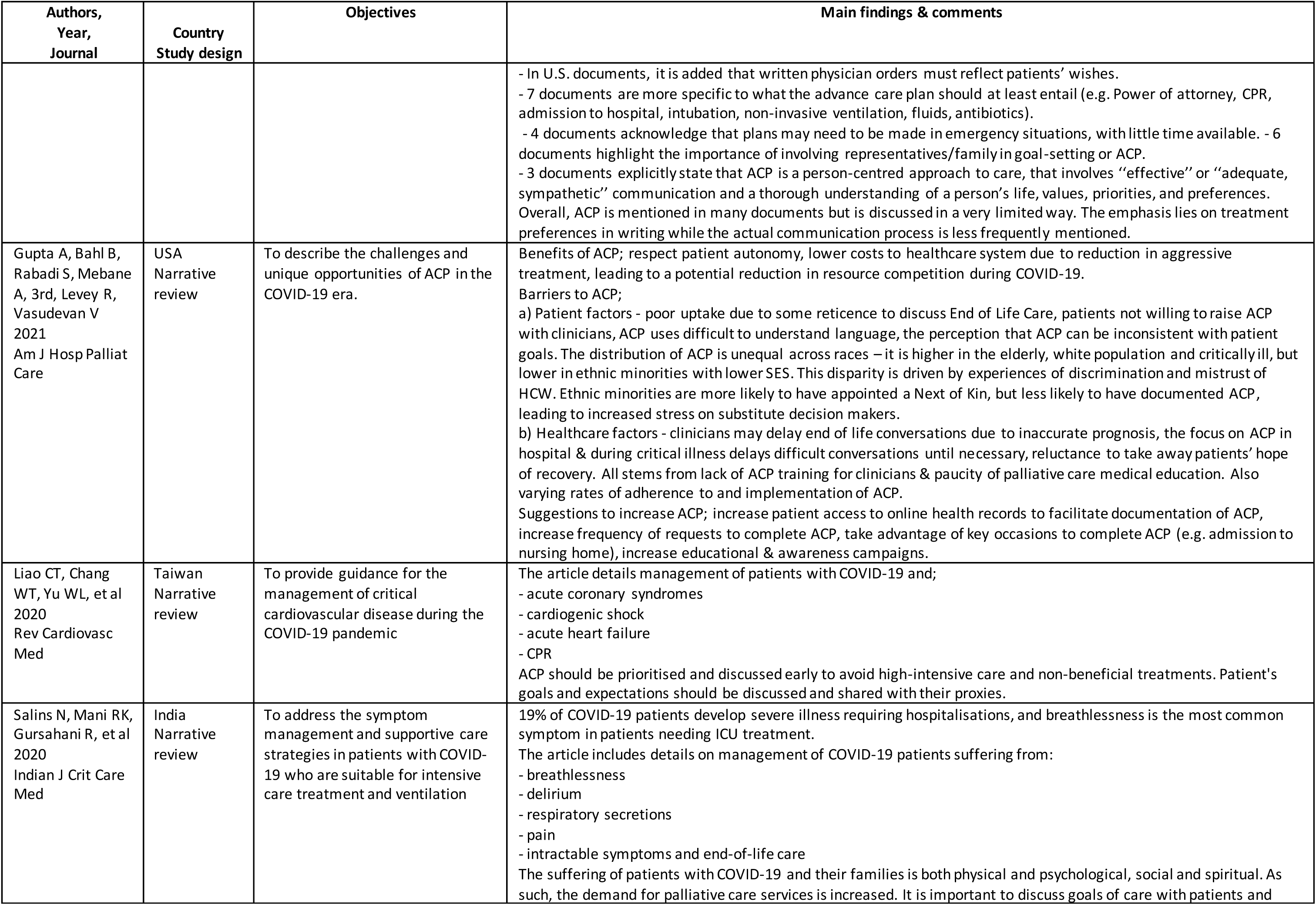
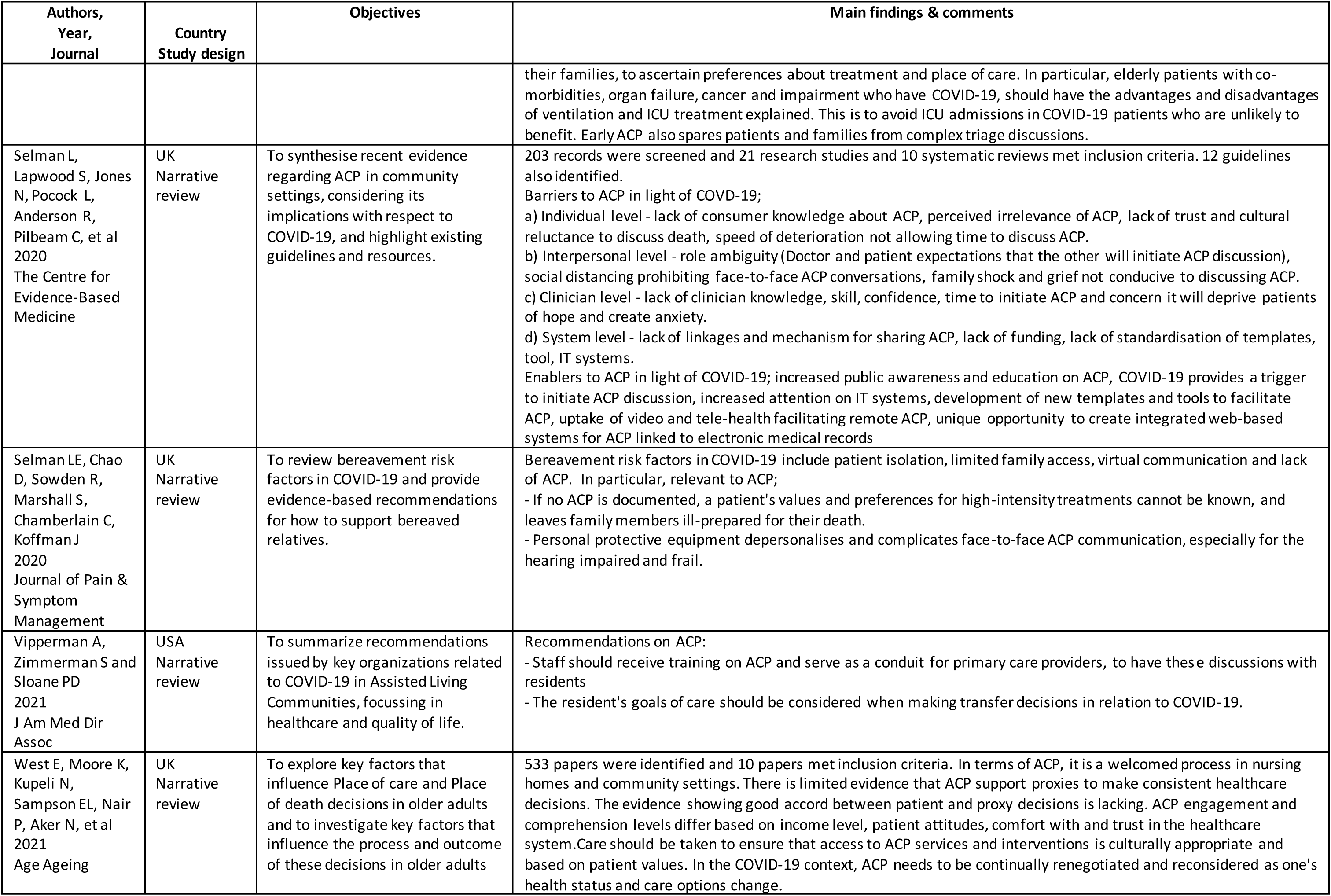
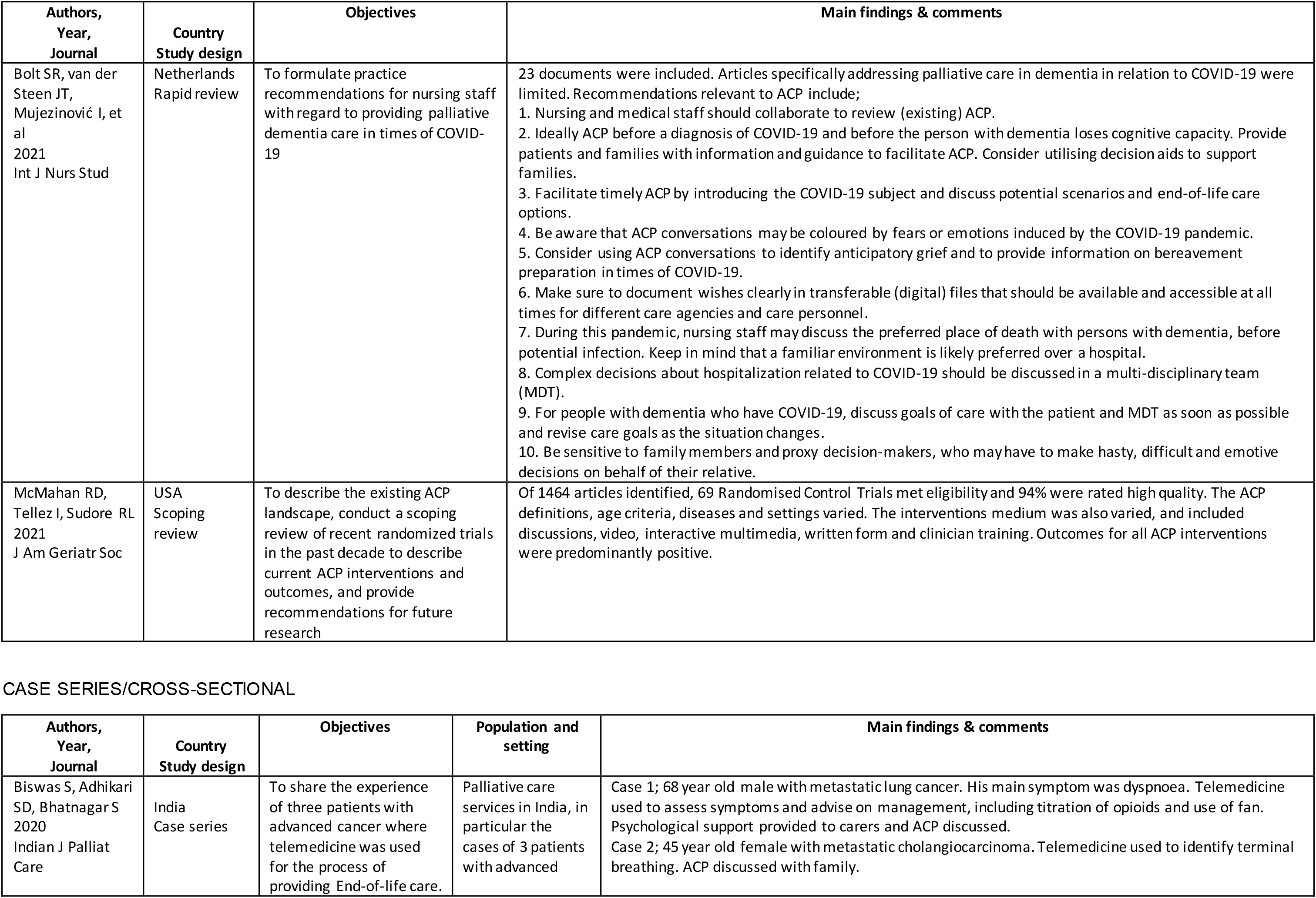
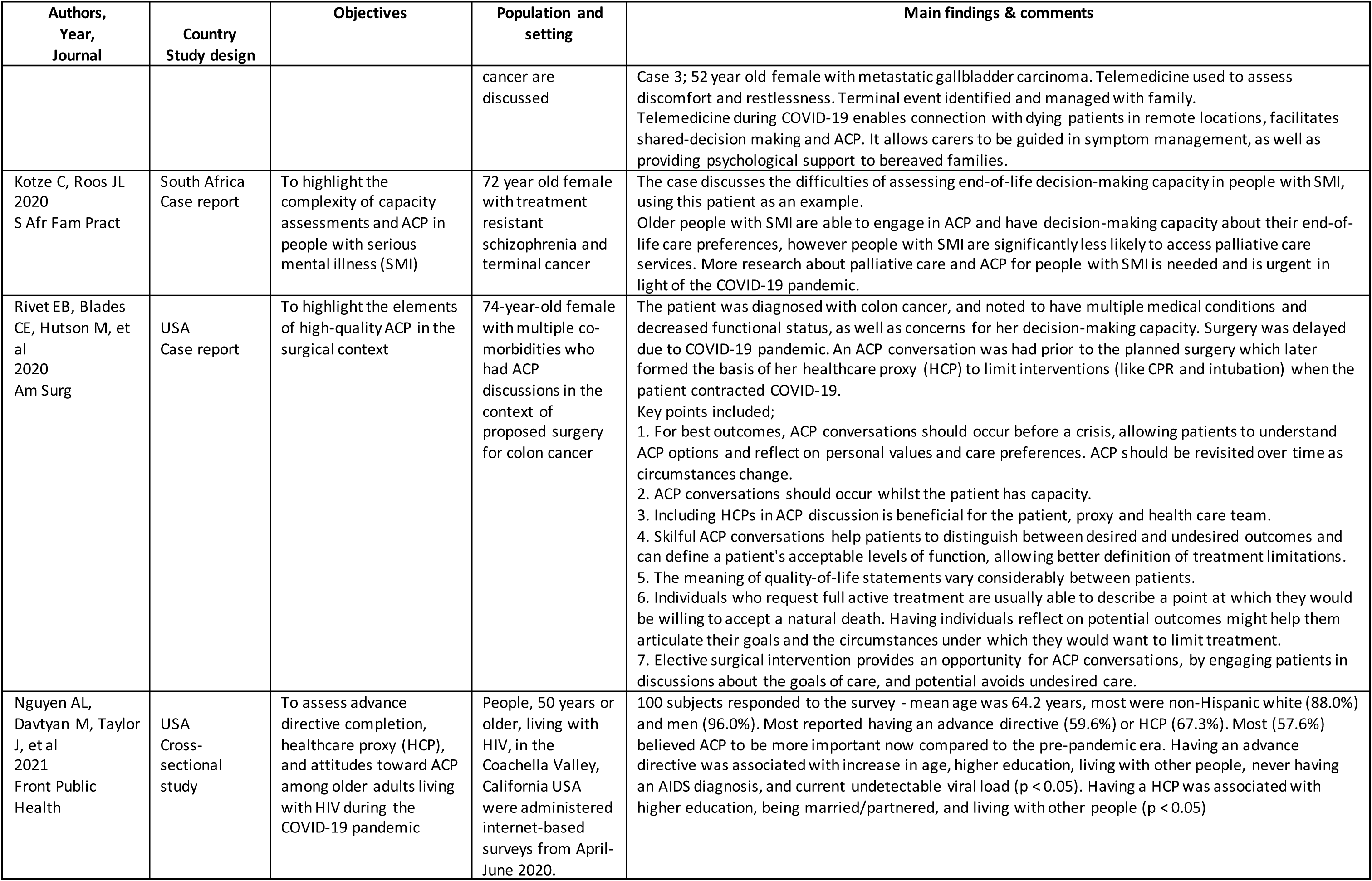
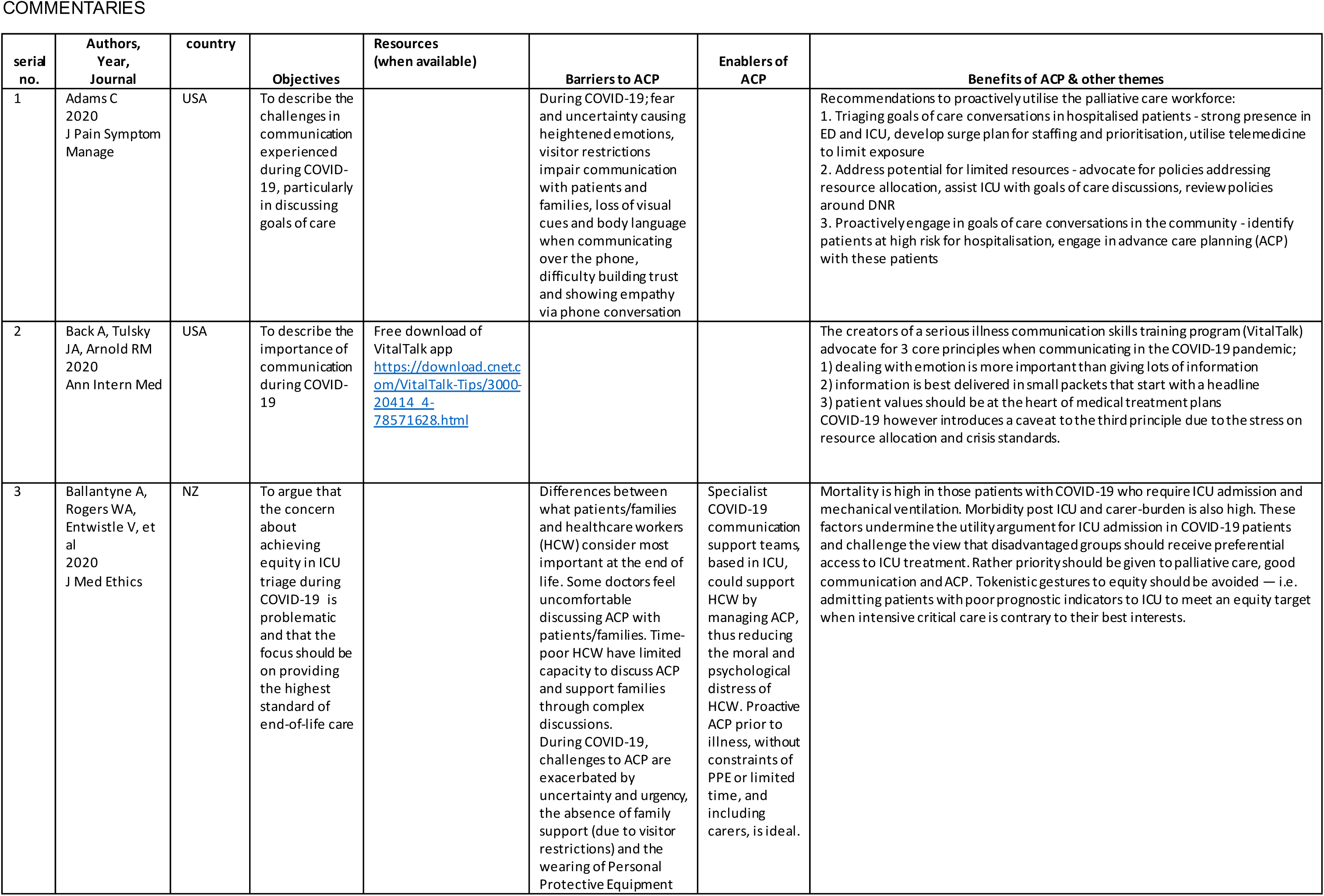
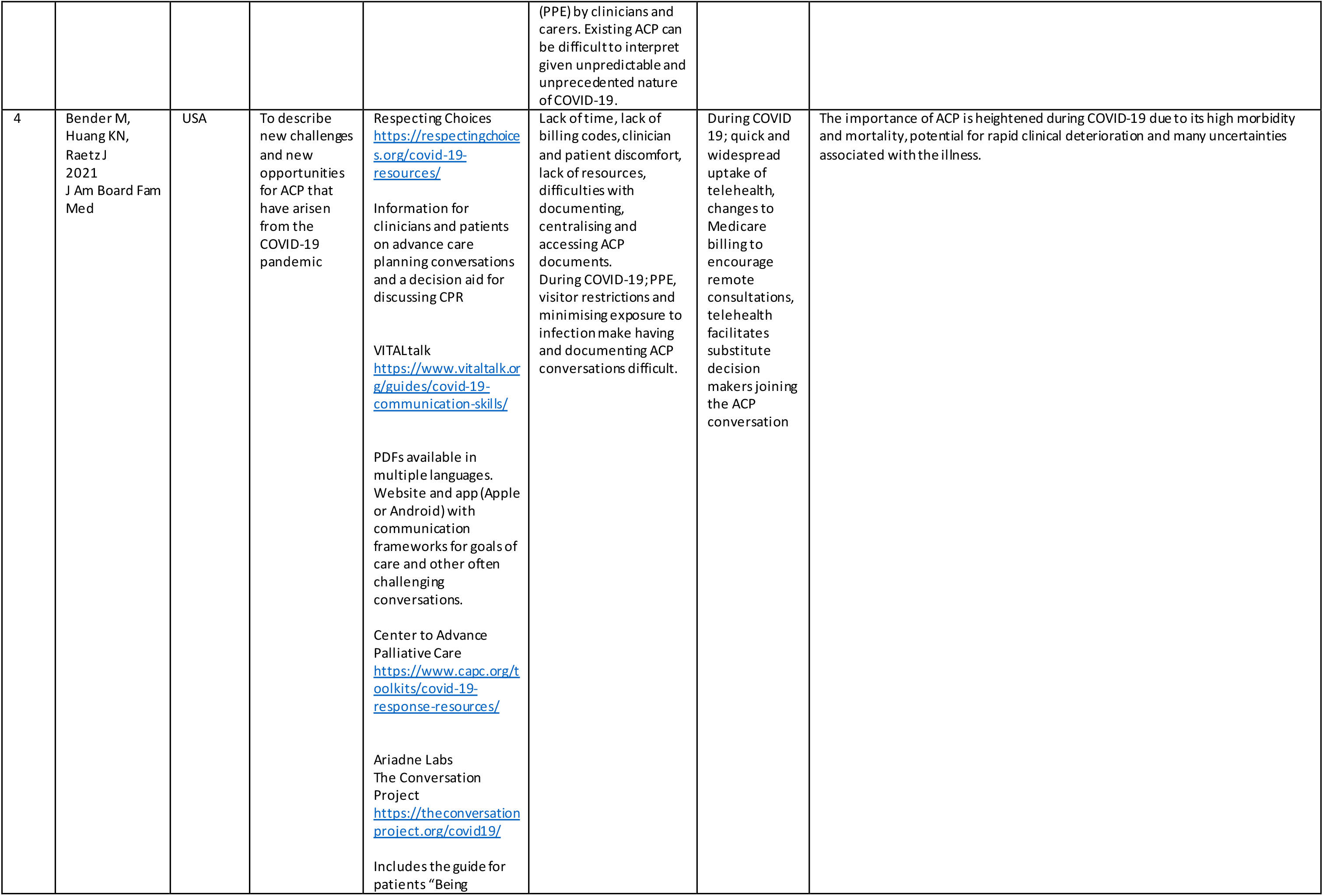
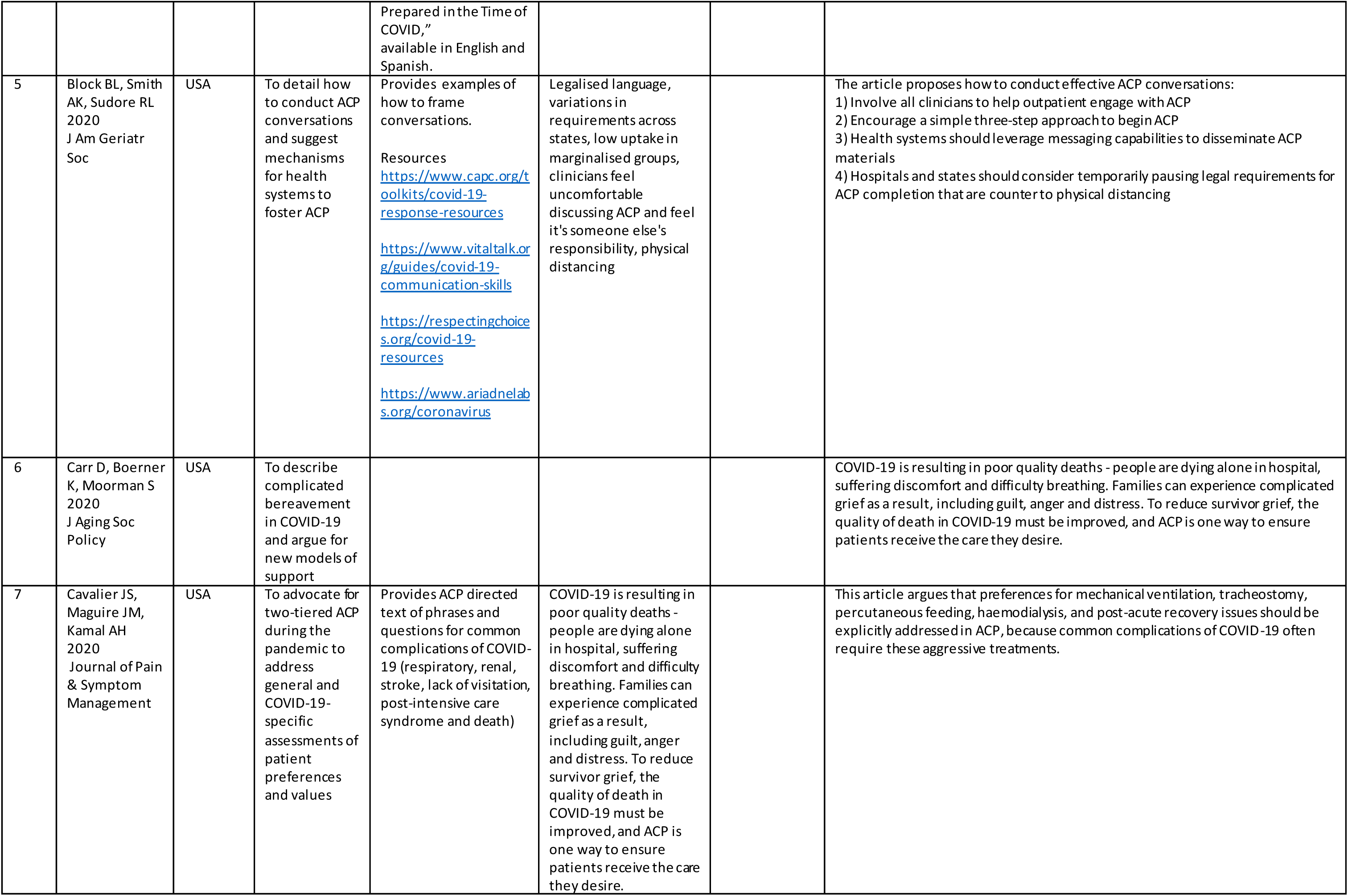
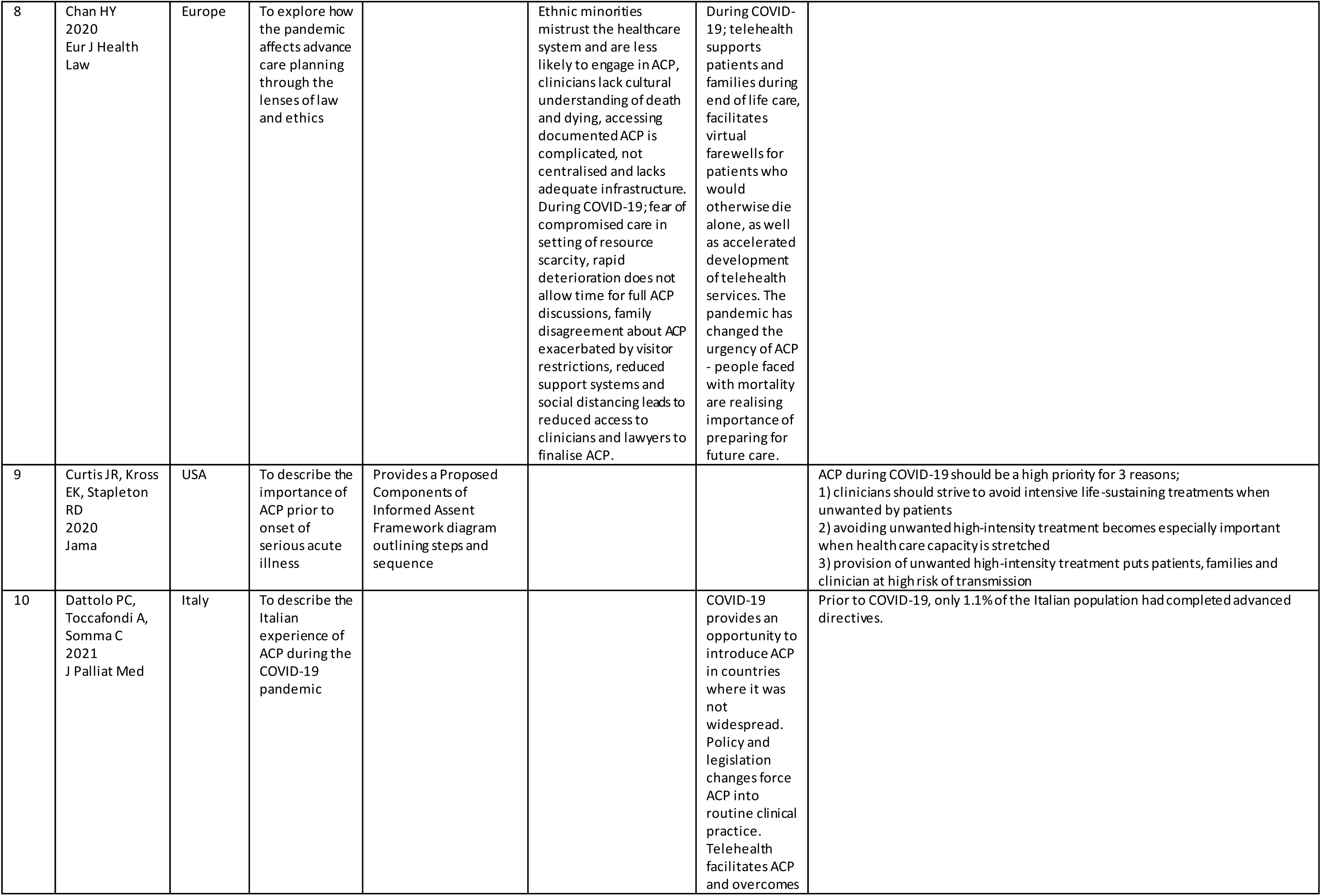
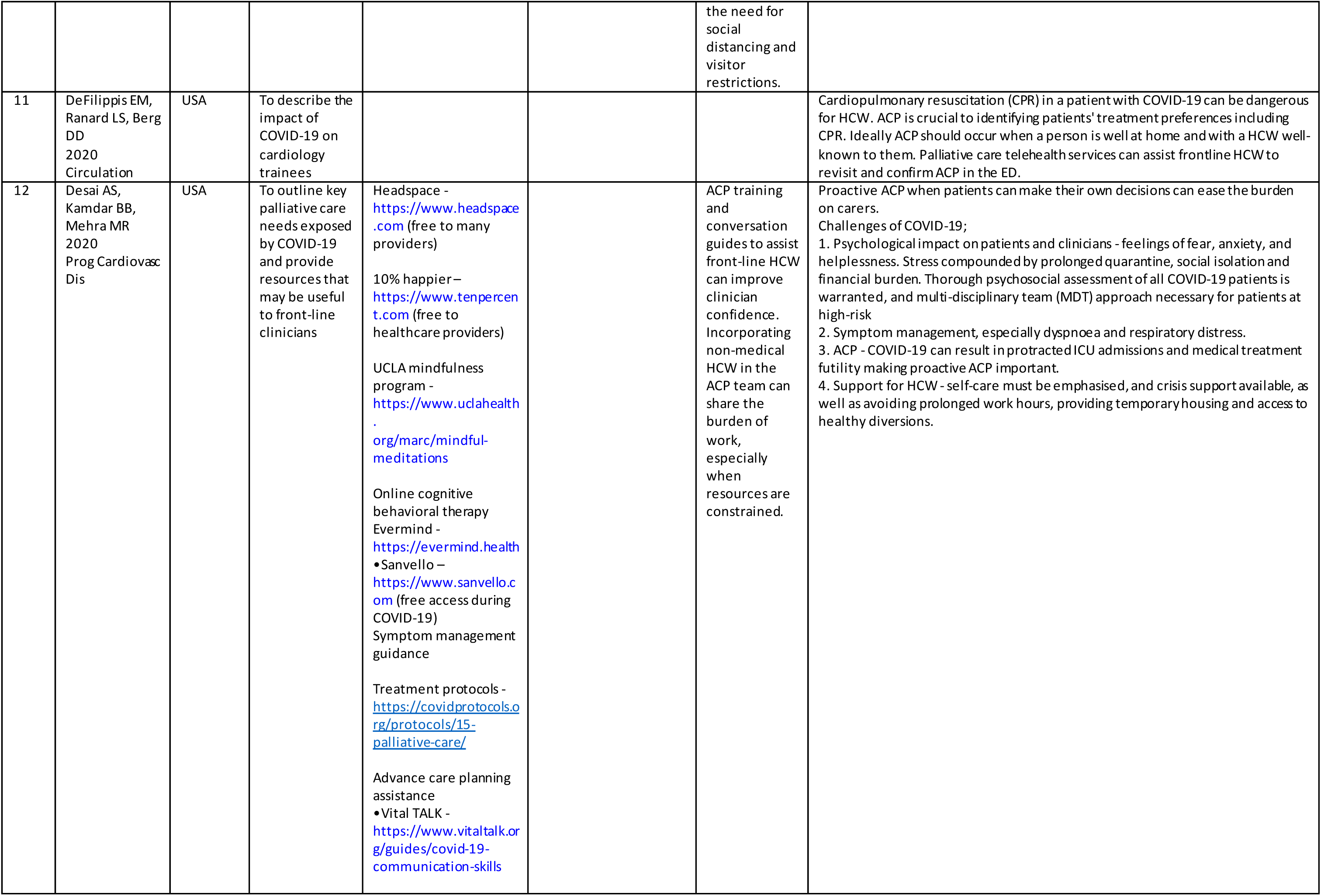
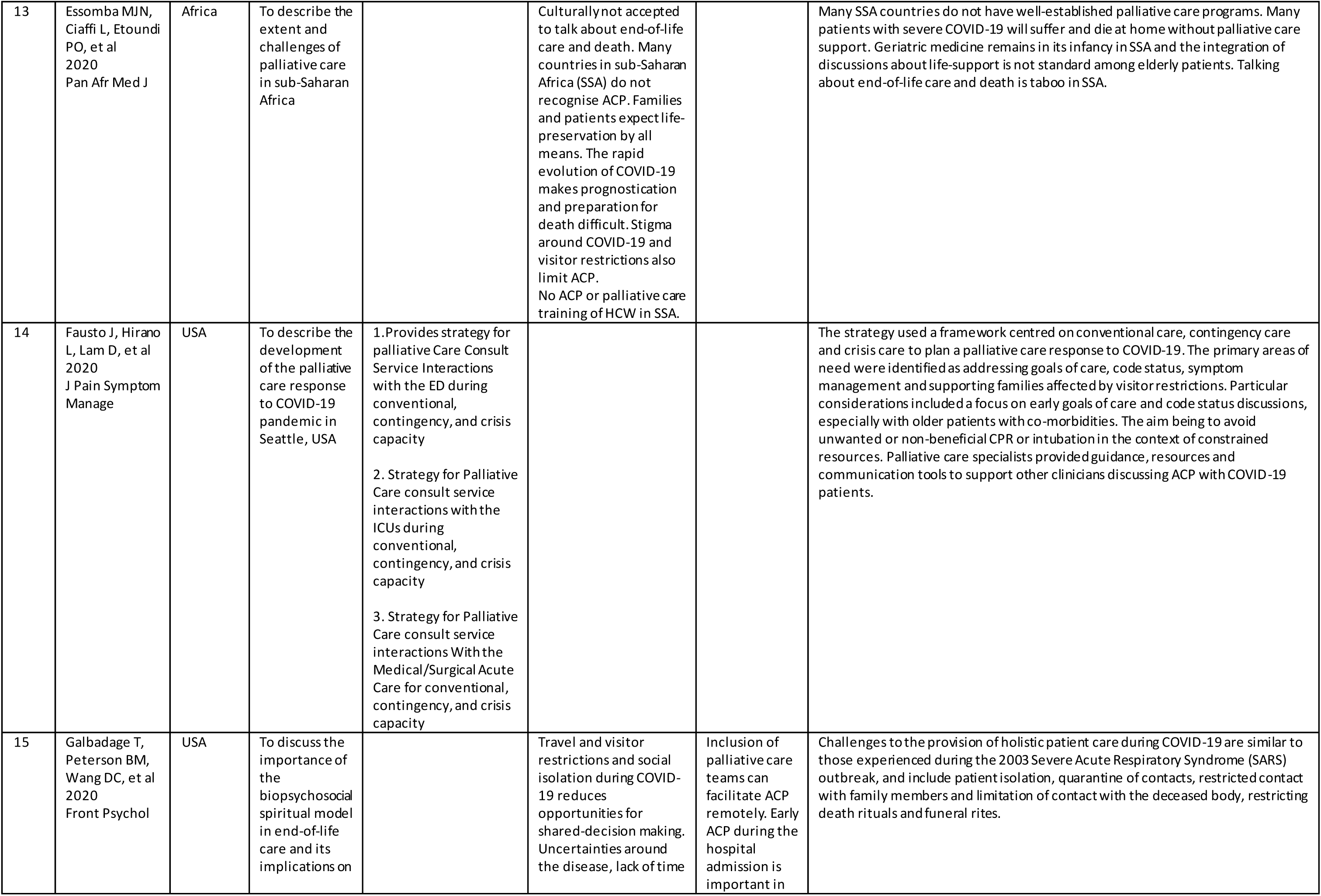
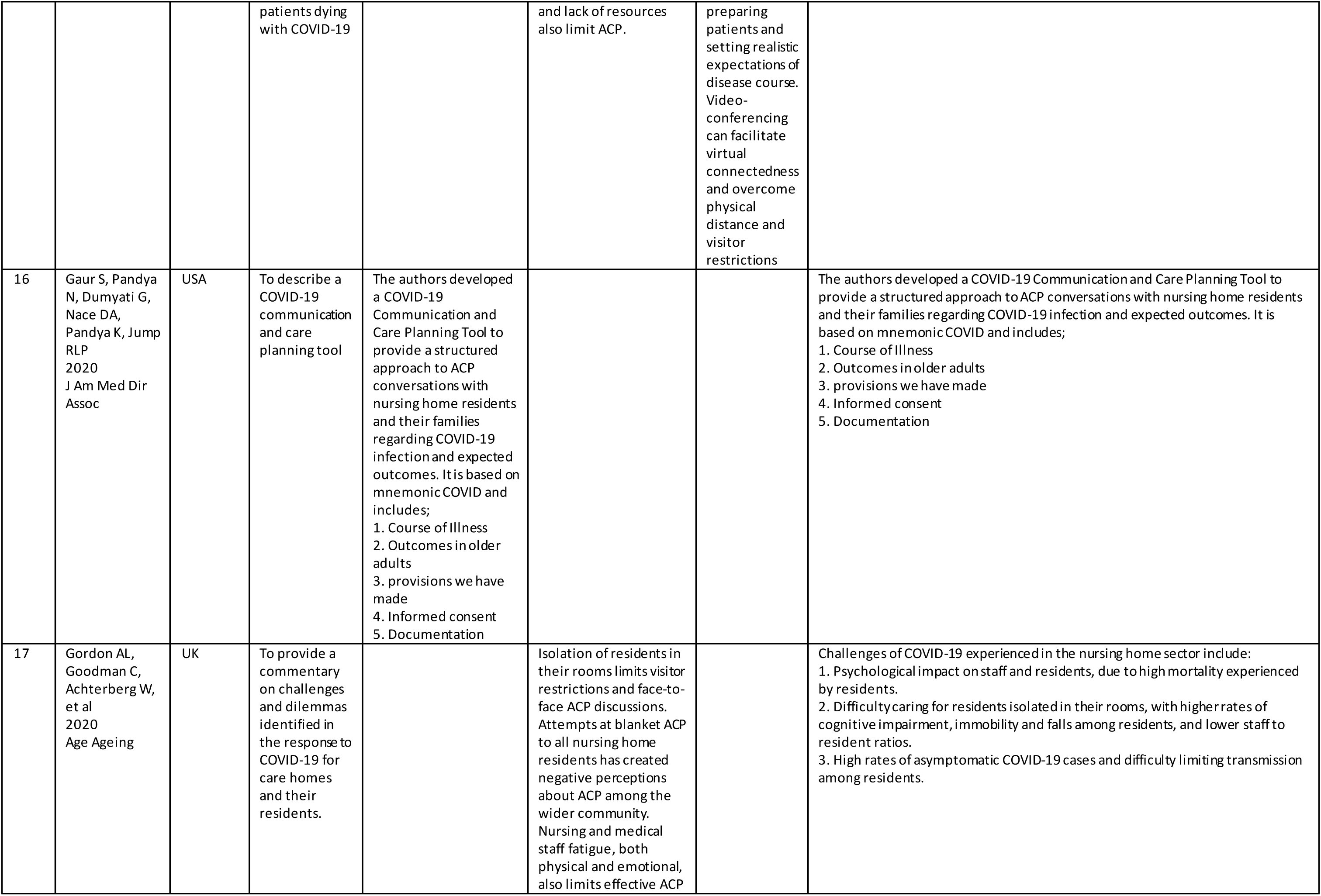
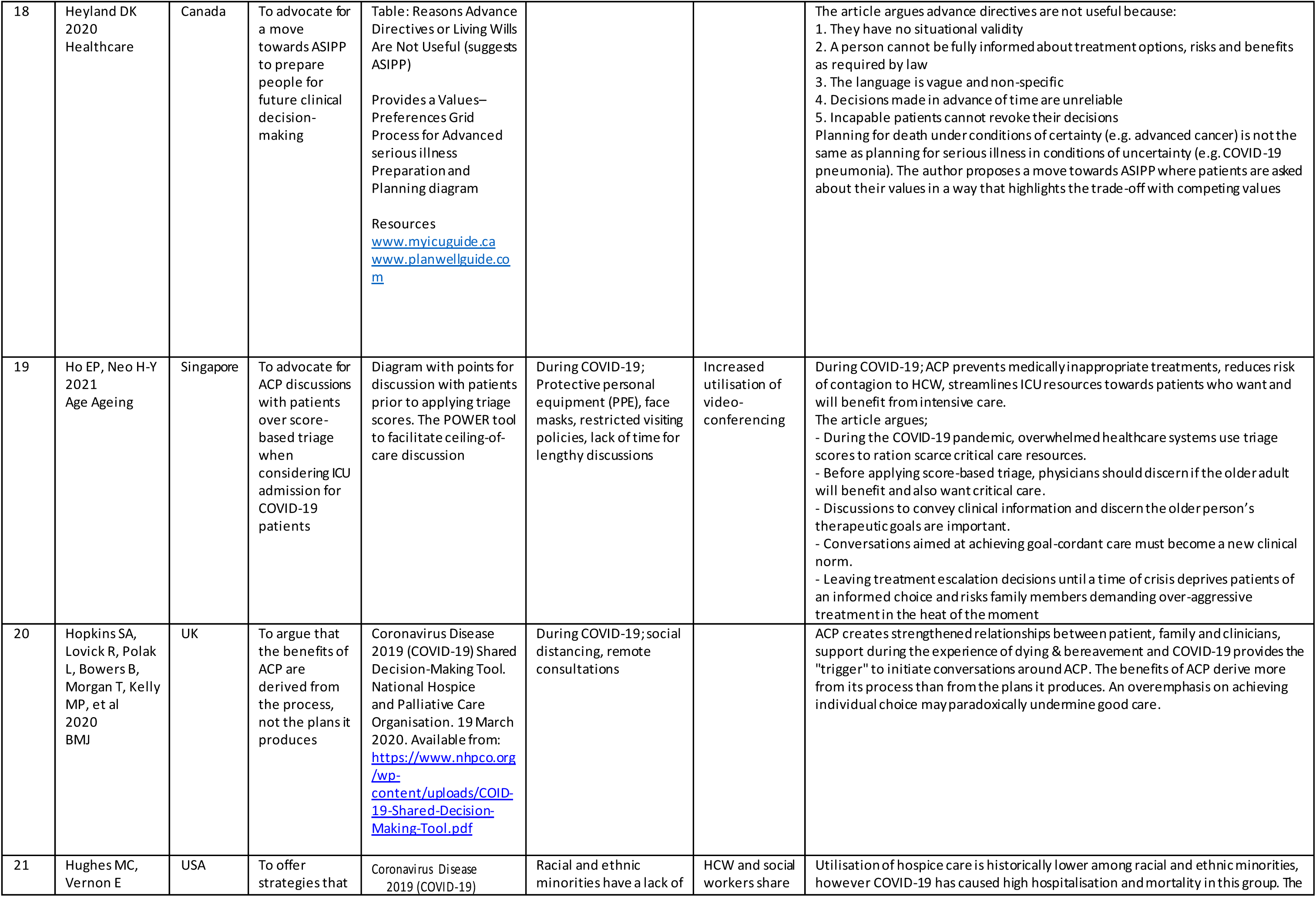
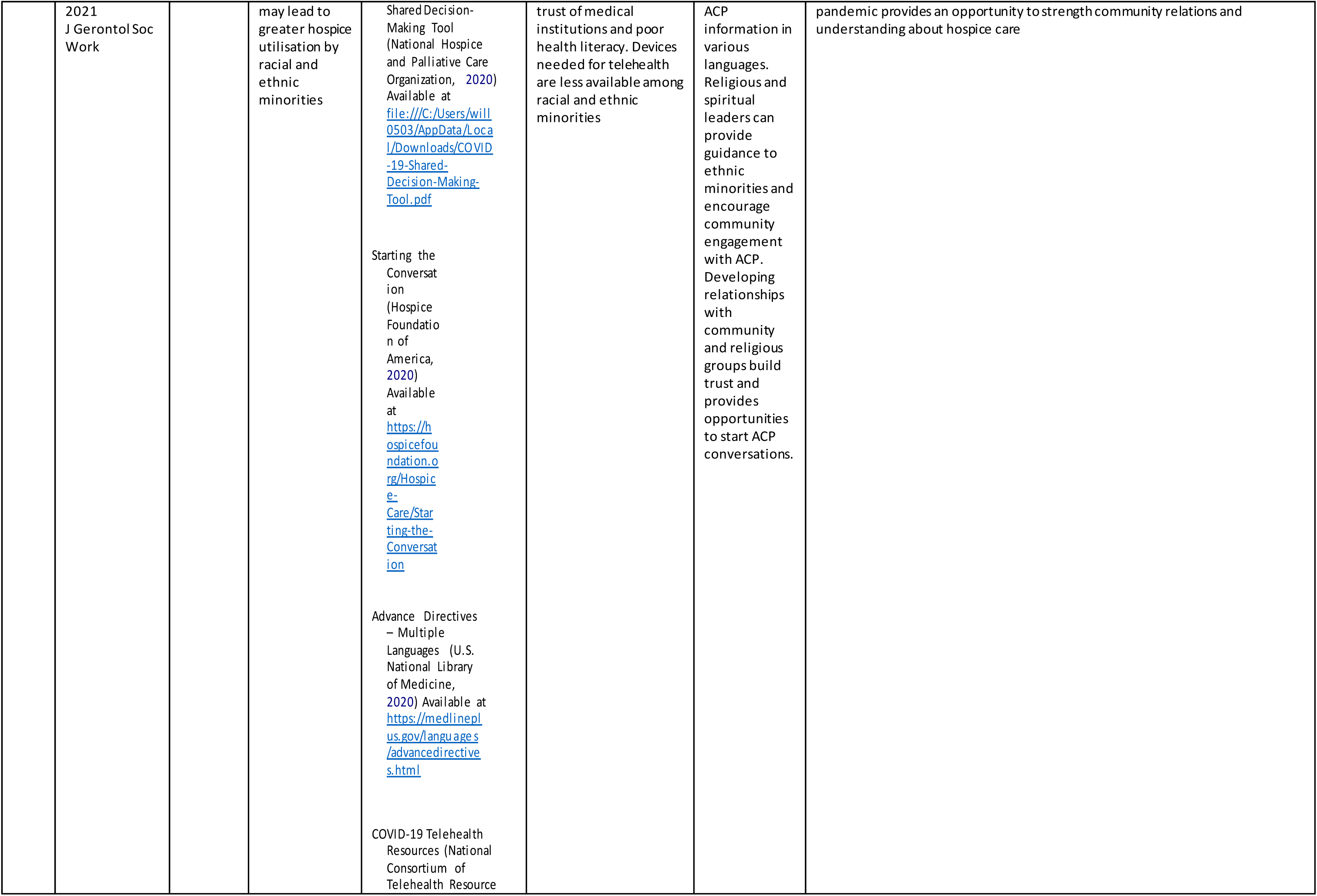
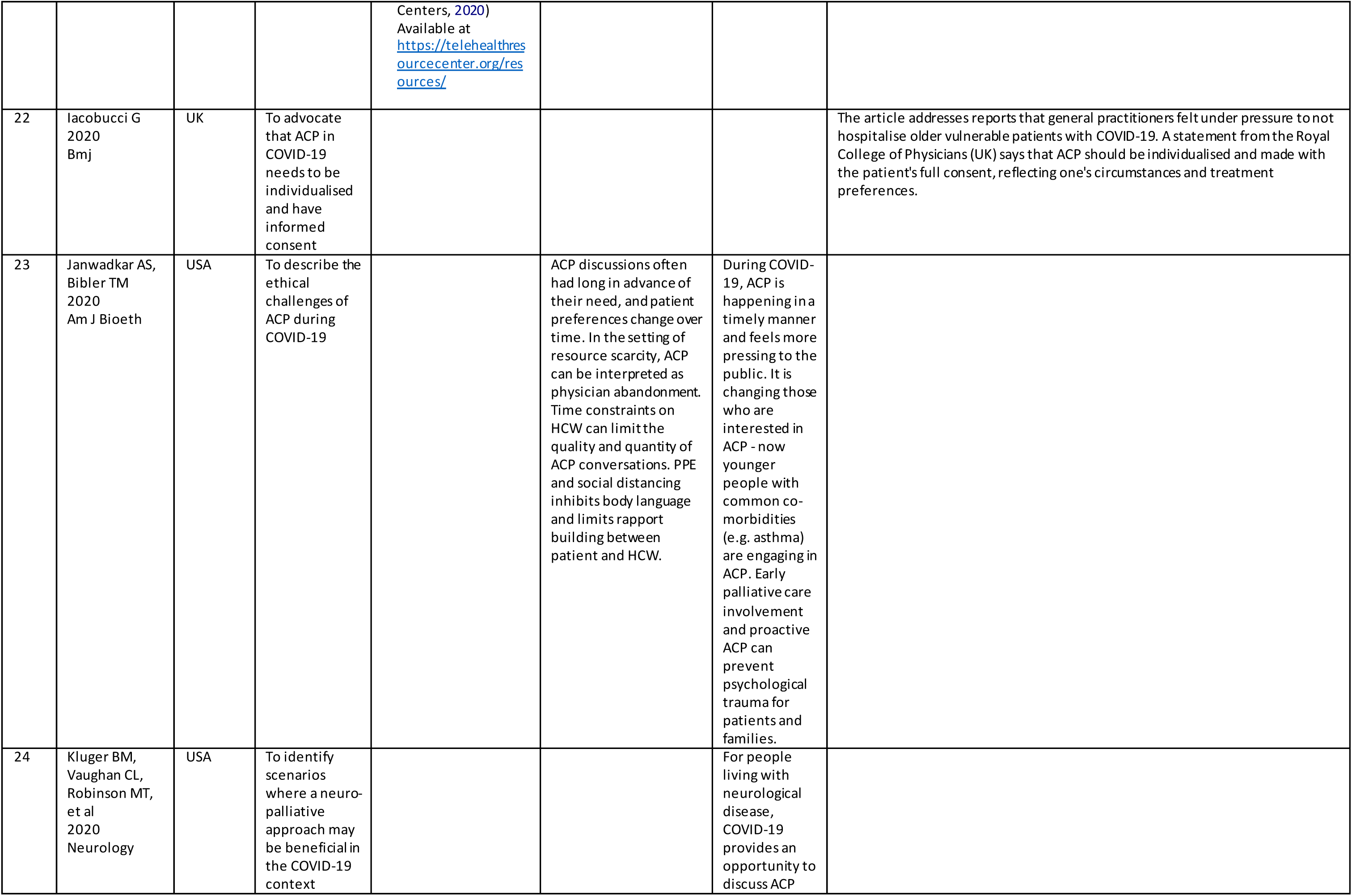
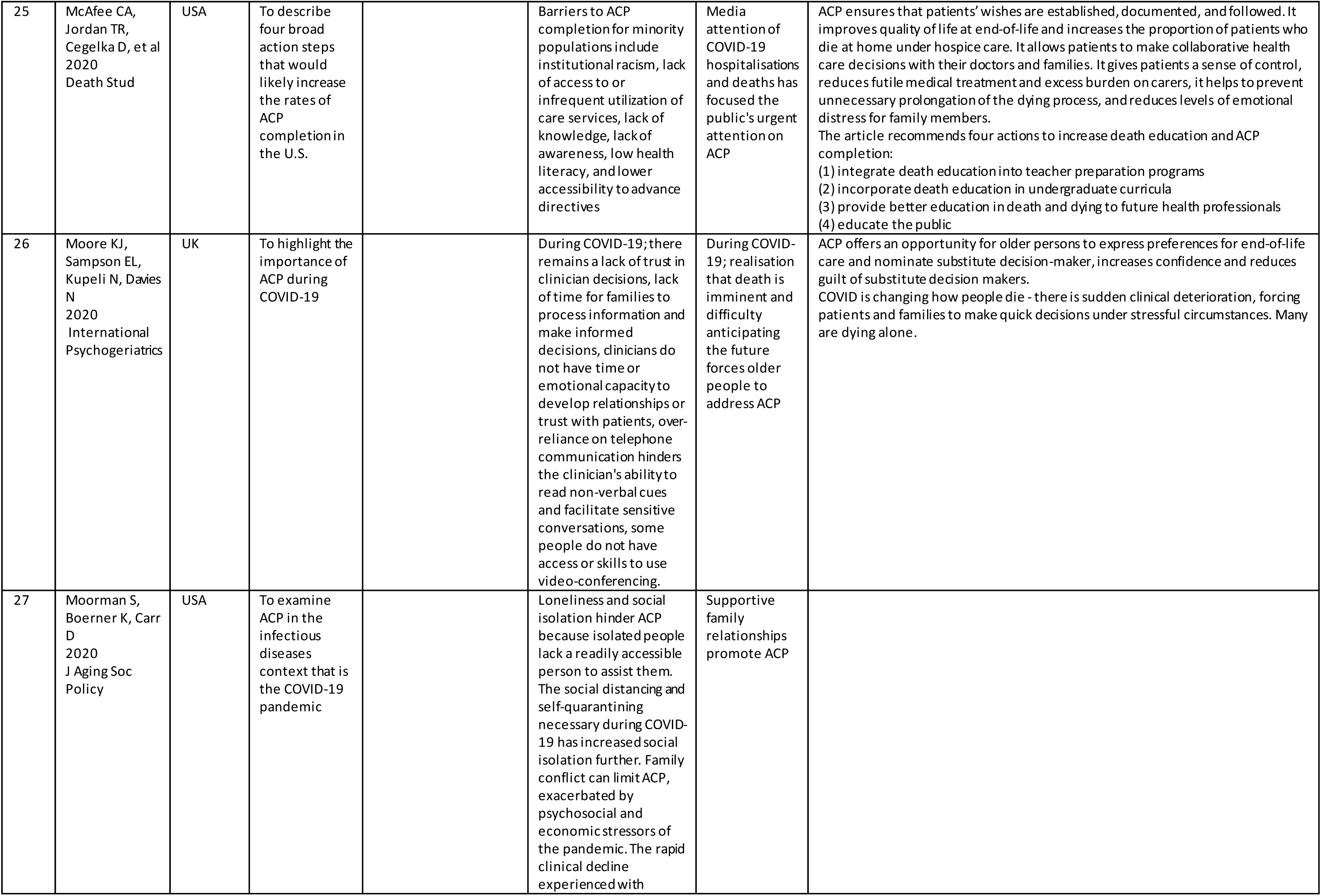
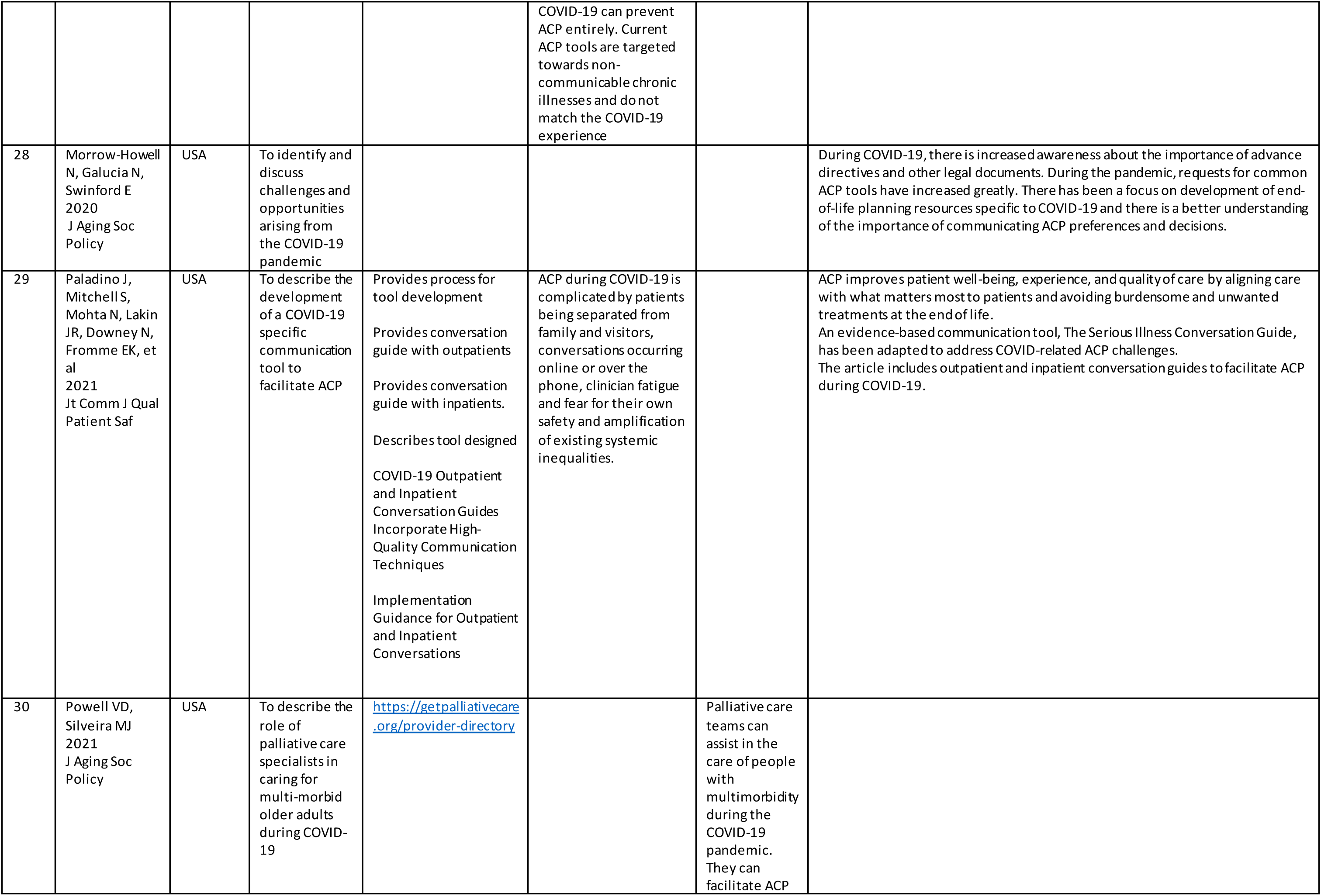
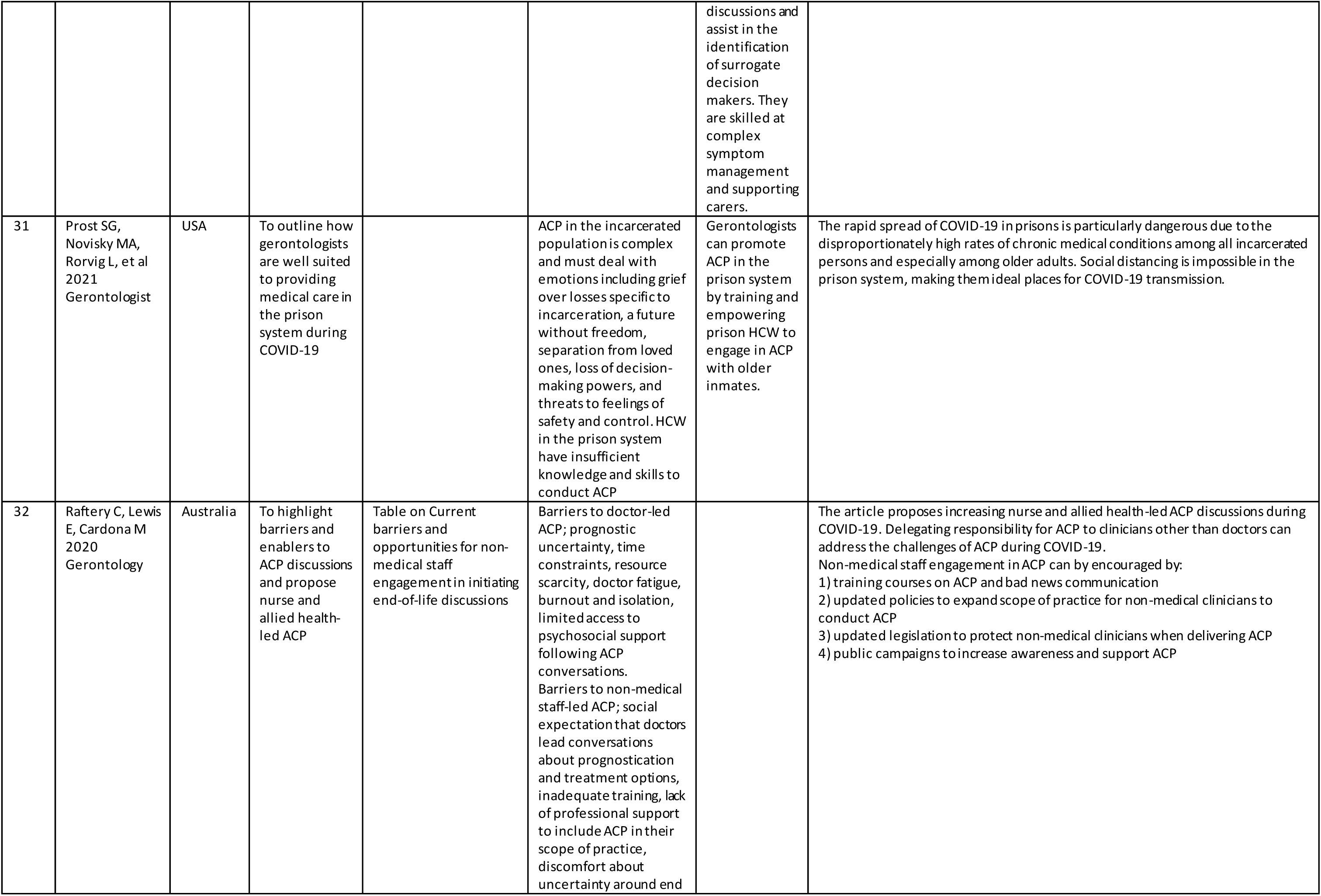
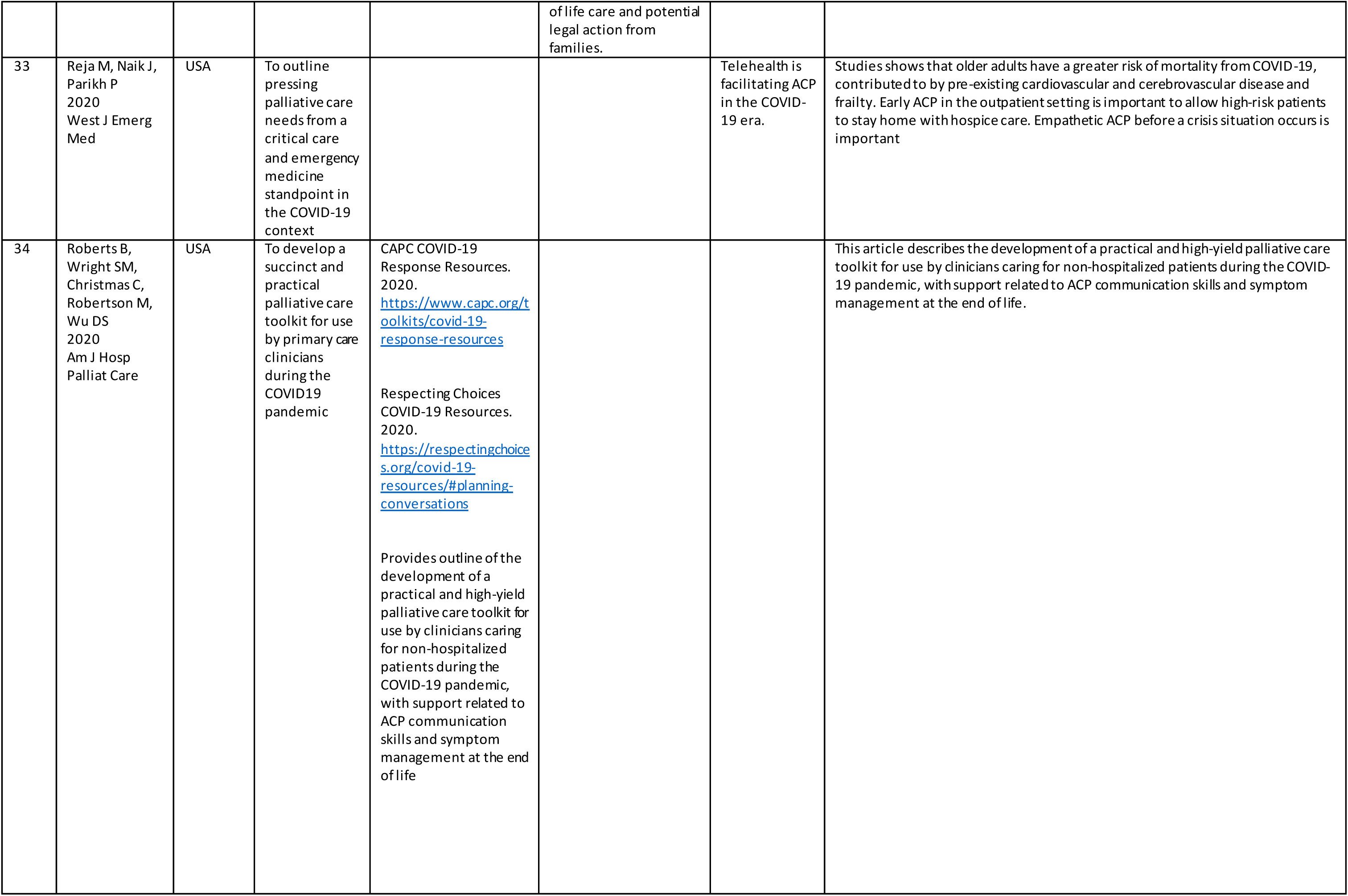
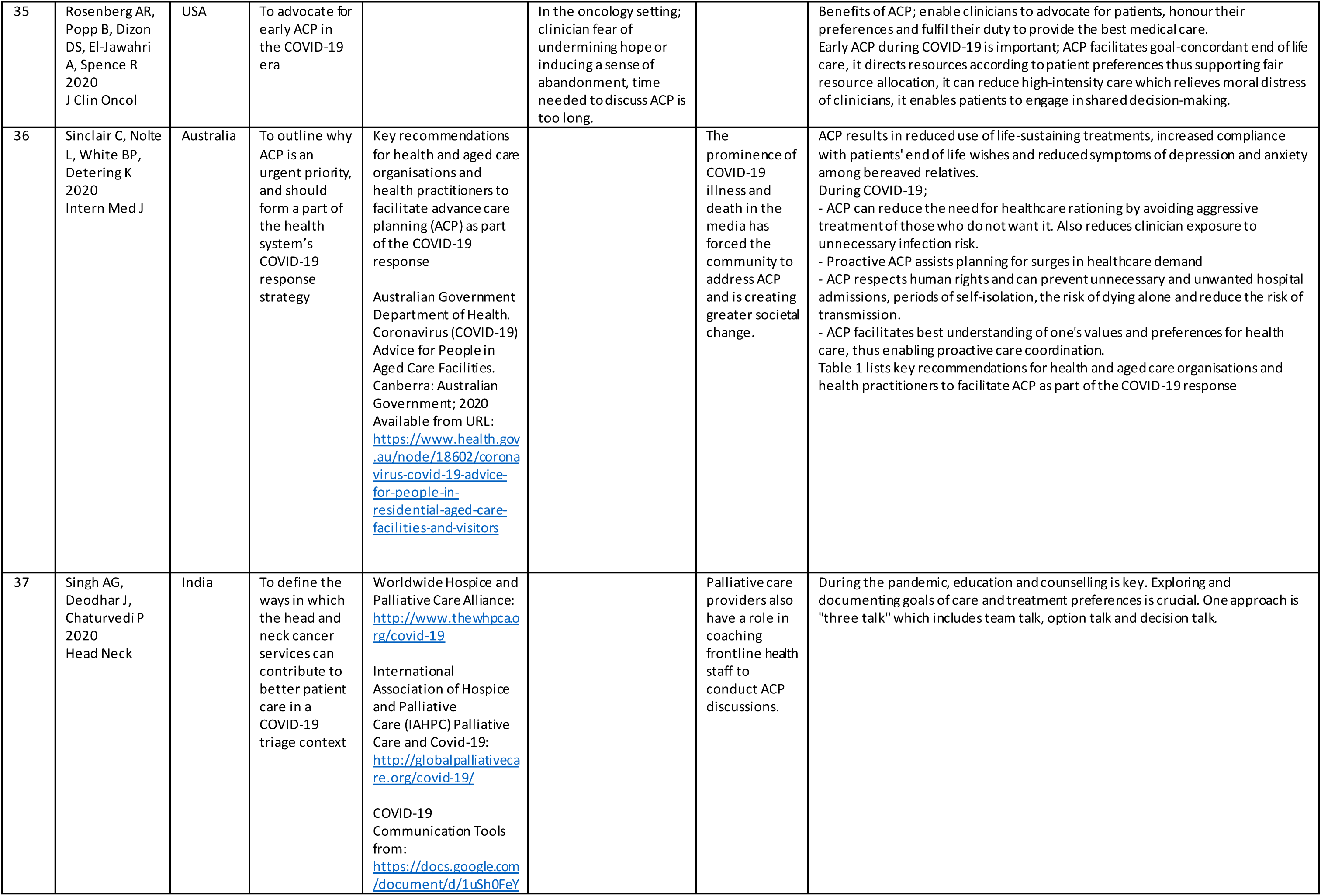
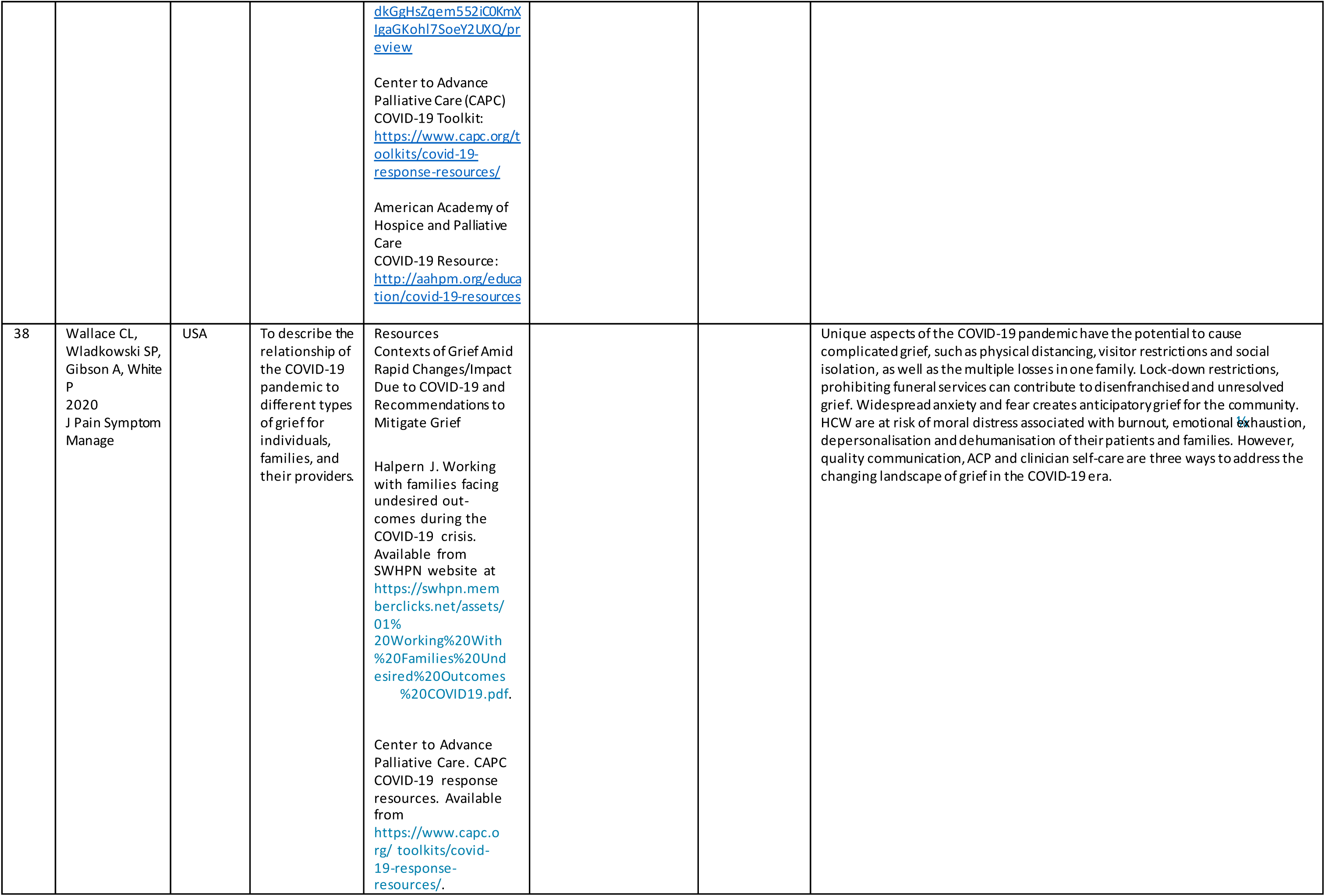
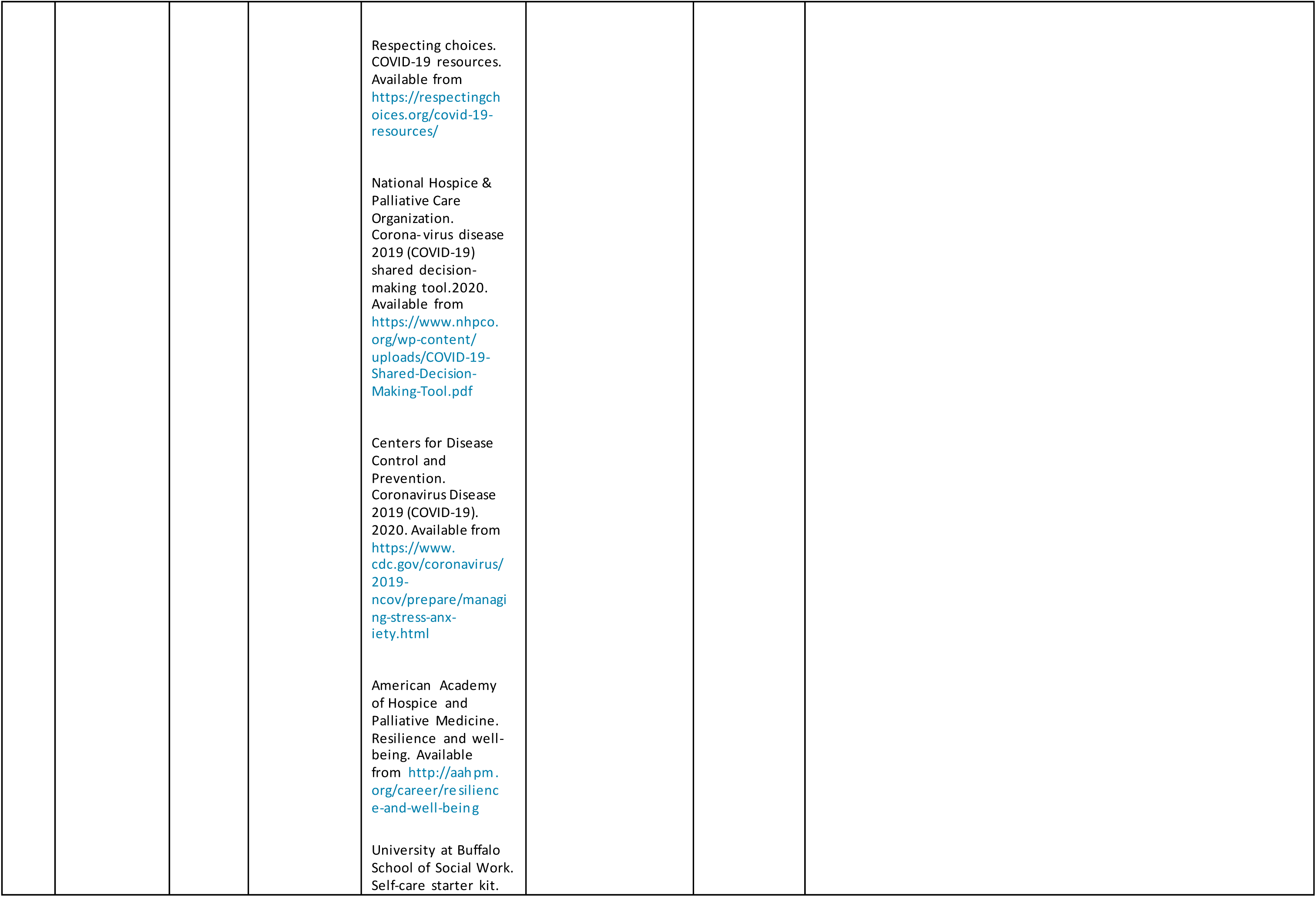
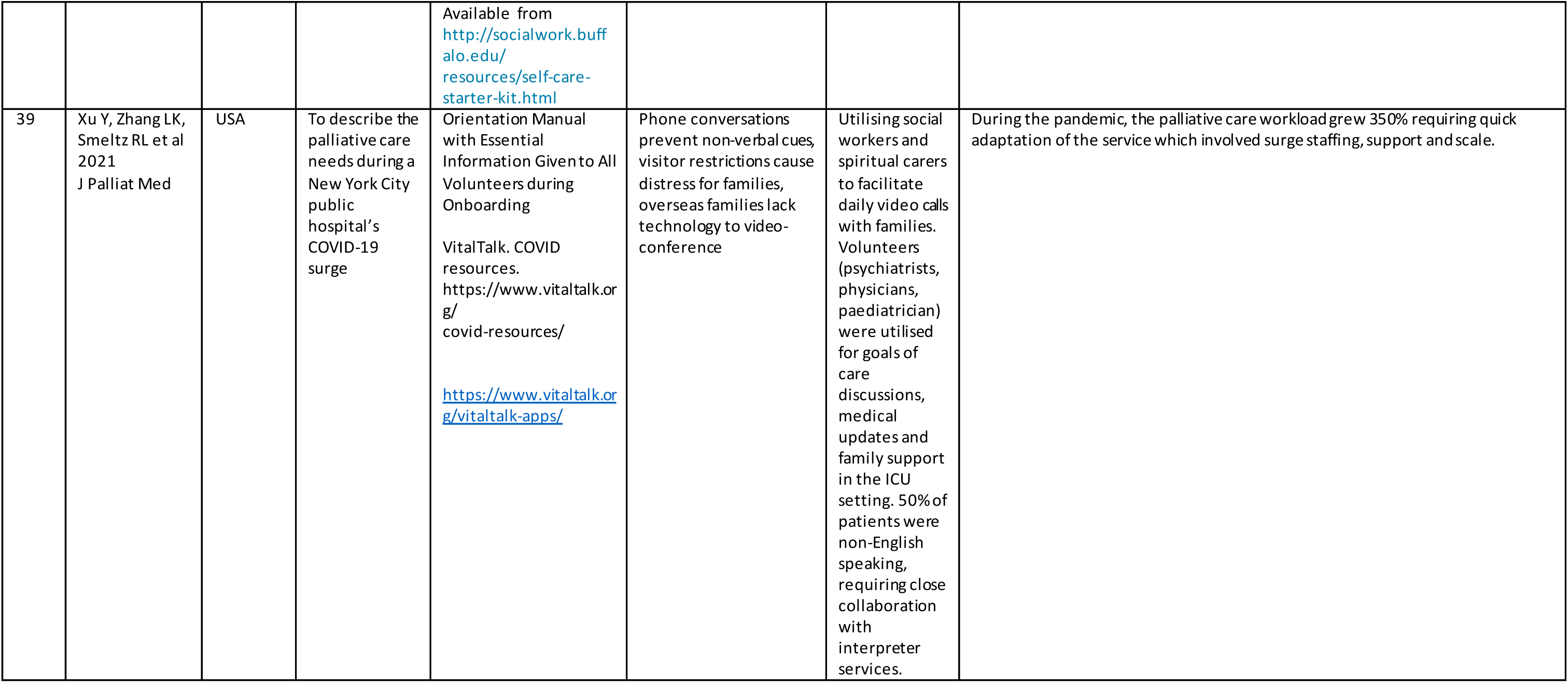
Supplement 1. Summary of Publications and Resources for Cohort Studies, Consensus Guidelines, Narrative Reviews, Cross-Sectional, Case Reports & Commentaries

## Notes

### Competing Interest Statement

The authors have declared no competing interest.

### Funding Statement

This study did not receive any funding

